# Single-cell multi-cohort dissection of the schizophrenia transcriptome

**DOI:** 10.1101/2022.08.31.22279406

**Authors:** W. Brad Ruzicka, Shahin Mohammadi, John F. Fullard, Jose Davila-Velderrain, Sivan Subburaju, Daniel Reed Tso, Makayla Hourihan, Shan Jiang, Hao-Chih Lee, Jaroslav Bendl, PsychENCODE Consortium, Georgios Voloudakis, Vahram Haroutunian, Gabriel E. Hoffman, Panos Roussos, Manolis Kellis

## Abstract

Schizophrenia is a prevalent mental illness with a high societal burden, complex pathophysiology, and diverse genetic and environmental etiology. Its complexity, polygenicity, and heterogeneity have hindered mechanistic elucidation and the search for new therapeutics. We present a single-cell dissection of schizophrenia-associated transcriptomic changes in the human prefrontal cortex across two independent cohorts, one deeply profiling 48 subjects (361,996 cells), and the other broadly profiling 92 subjects (106,761 cells). We identified 25 cell types that we used to produce a high-resolution atlas of schizophrenia-altered genes and pathways. Excitatory neurons were the most affected cell group, with transcriptional changes converging on neurodevelopment and synapse-related molecular pathways. Differentially expressed gene sets implicate a coherently expressed module of trans-acting regulatory factors involved in neurodevelopment and genetically associated with schizophrenia risk. Transcriptional alterations significantly overlapped with known genetic risk factors, suggesting convergence of rare and common genomic variants on reproducible neuronal population specific alterations in schizophrenia. The severity of transcriptional pathology segregated two populations of schizophrenia subjects in a manner consistent with the expression of specific transcriptional patterns marked by genes involved in synaptic function and chromatin dynamics. Our results provide a high-resolution single cell atlas linking transcriptomic changes within specific cell populations to etiological genetic risk factors, contextualizing established knowledge within the cytoarchitecture of the human cortex and facilitating mechanistic understanding of schizophrenia pathophysiology and heterogeneity.

## Introduction

Schizophrenia is a neuropsychiatric disorder clinically characterized by a combination of psychosis, social withdrawal, and cognitive dysfunction, often leading to a lifetime of profound and chronic disability (1). Its pathogenesis is thought to begin during neurodevelopment, yet first psychotic episodes do not occur until early adulthood (2). The complex etiology of schizophrenia involves genetic and environmental factors presumed to affect a wide range of brain-related processes, including neurodevelopment (3, 4), synaptic function (5, 6), neuronal excitability (7, 8), neuronal connectivity (9, 10), and cognition (11, 12). Despite substantial progress in schizophrenia genetics (13–15) and the functional genomic examination of postmortem tissue (16–23), elucidating specific molecular and cellular alterations linking etiological risk factors and clinical presentation remains challenging.

Cell-type-specific molecular changes in schizophrenia have been previously reported using targeted methods such as laser microdissection (16–23) and fluorescence-activated cell sorting (24–26), identifying changes within major neuronal classes pointing to the selective transcriptional vulnerability of deep-layer excitatory and parvalbumin-expressing interneurons. While these approaches have revealed much about schizophrenia molecular pathophysiology within the context of human brain cytoarchitecture, limitations include their reliance on marker gene expression to select cells of interest, limited ability to dissect subpopulations of major cell-types, and the preselection of target cell-types. Emerging technologies for massively parallel single-cell transcriptomics (27, 28) achieve cell-type-specificity without bias toward preselected markers and populations, enabling the discovery of disease-associated changes as recently demonstrated for Alzheimer’s Disease (29), Autism Spectrum Disorder (30), Major Depressive Disorder (31), and Multiple Sclerosis (32).

To investigate which cell types within the complex cytoarchitecture of the human brain present reproducible expression changes associated with schizophrenia, we profiled postmortem prefrontal cortex tissue samples from two independent cohorts using single-nucleus RNA sequencing (snRNA-seq) (**Fig. 1a**). The McLean (McL) cohort included 24 schizophrenia and 24 control subjects, balanced for sex (12 male and 12 female subjects per group), age (ranging from 22 to 94 years, average 63.5 years), and postmortem interval (ranging from 6.9 to 26.3 hours, average 16.8 hours). The Mount Sinai School of Medicine (MSSM) cohort included 41 schizophrenia and 51 control subjects, balanced for age (ranging from 24 to 101 years, average 72.7 years) and postmortem interval (ranging from 2.0 to 52.8 hours, average 18.1 hours), including both male (61) and female (31) subjects (Supplementary Table 1). In order to increase the number of cells captured from each individual while reducing batch effects, we implemented a multiplexing strategy pooling a mixture of cases and controls in each sequencing library (Methods). We obtained and report a total of 468,727 high-quality single-nucleus transcriptomes, including 206,014 nuclei from schizophrenia and 262,713 from control individuals, profiled at an average depth of 12k cells per subject and 35k reads per cell (McL) or 1.2k cells per subject and 58k reads per cell (MSSM). We used these data for the remainder of the study (Supplementary Table 2).

**Figure 1.**
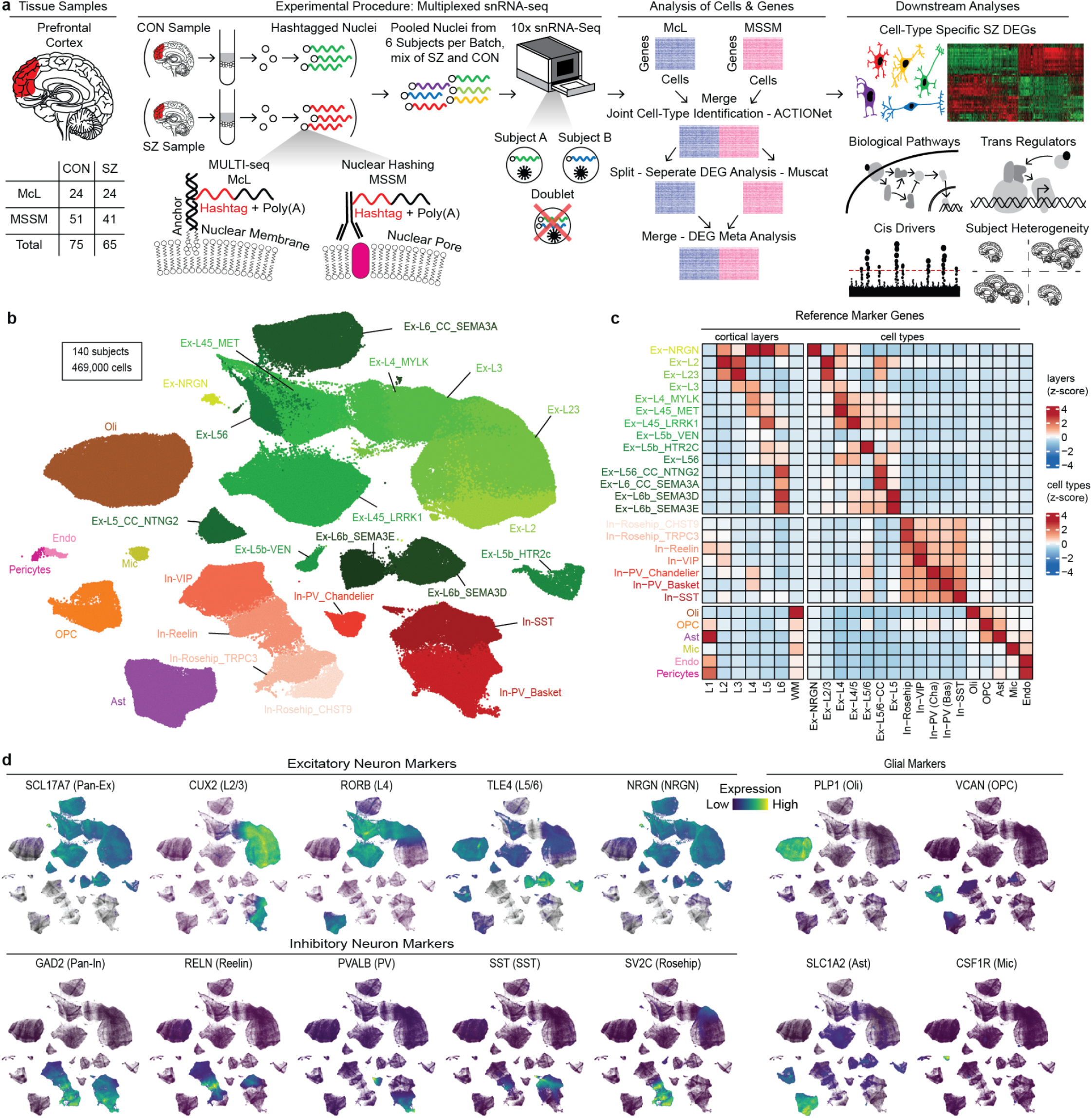
Multiresolution dissection of cellular subpopulations. **a.** Overview of study design and data analysis strategies, depicting pooling of nuclei after sample-specific hashtag labeling allowing removal of intersample doublets from the data, and analysis strategy of pooling datasets for identification and annotation of cell types before DEG analysis within each dataset independently and merging of results through meta-analysis. On the right downstream analyses of biological pathways and cis and trans-regulatory factors related to cell type-specific schizophrenia DEG sets are depicted. **b.** ACTIONet plot of putative cell types. Green and red clusters represent excitatory and inhibitory subtypes of neurons respectively, with darker shades indicating an association with deeper cortical layers. **c.** Annotation of cell types using curated markers(33) from previous single-nucleus RNA-seq (29, 30, 39) and spatial transcriptomics (40) studies. **d.** Projection of known marker genes verifies cell type annotations and cortical layer associations (41).

### Identification of cellular populations across cohorts

We identified major brain cell types and neuronal subpopulations across both cohorts by building and annotating a cell similarity network using ACTIONet (33). Major brain cell classes including excitatory and inhibitory neurons, astrocytes, oligodendrocytes, oligodendrocyte progenitor cells, and microglia were identified by preferential expression of known marker genes. By extracting and clustering each major cell class, we identified additional low-abundance cell types and neuronal subpopulations for a total of 27 cell types and subpopulations capturing all major cell classes of the human prefrontal cortex, including excitatory and inhibitory neuron subtypes and glial cells, with cell types well represented across samples and cohorts (**Fig. 1b, Extended Data Fig. 1a-h**). Excitatory neuronal subpopulations preferentially expressed cortical layer marker genes and were annotated accordingly (**Fig. 1c**, Supplementary Table 3). All annotations were consistent with expression patterns of selected marker genes across the cell similarity network (**Fig. 1d**). Neuronal subtypes, and excitatory neurons in particular, had higher numbers of expressed genes and unique molecular identifiers (UMIs) (**Extended Data Fig. 2**), consistent with previous observations (29, 30).

**Figure 2.**
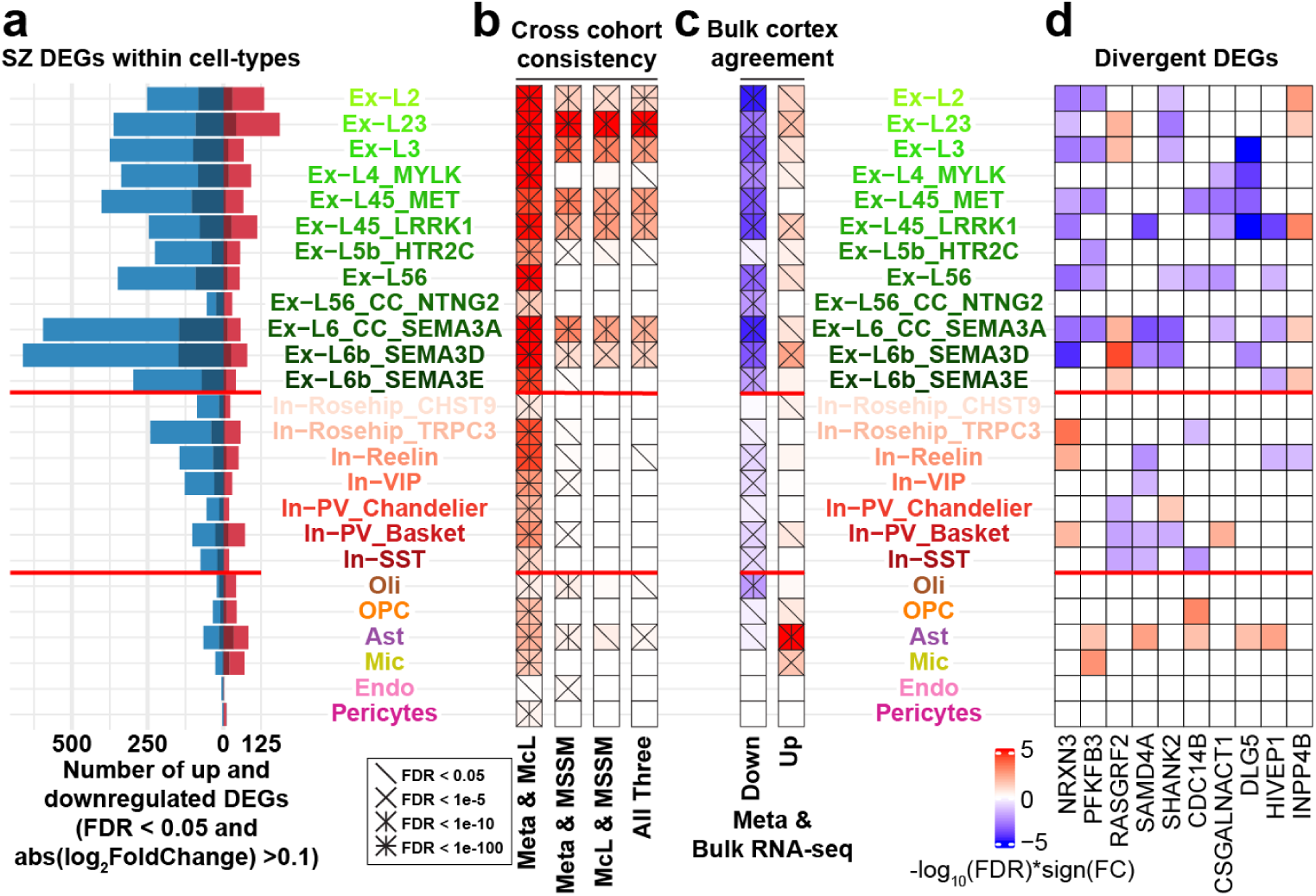
Cell-type-specific differential gene expression in schizophrenia. **a.** Barplots indicate the number of down (blue) and upregulated (red) genes with FDR < 0.05 and abs(log_2_(fold change) > 0.1 in meta-analysis combining results of both cohorts. Shaded regions of each bar indicate the number of genes also significantly different with the same direction of change in a bulk tissue RNA-seq study of schizophrenia prefrontal cortex (42). **b.** Heatmaps depict the significance of the overlap of cell type-specific DEG sets identified in each independent cohort and in the meta-analysis. Columns are scaled independently and cross hatches indicate the significance of each tested overlap. **c.** Concordance of meta-analysis DEGs and DEGs identified by prior bulk cortex RNA-sequencing (42) with up and downregulated DEGs considered separately. As in panel b, columns are scaled independently and cross hatches indicate the significance of each tested overlap. **d.** Signed significance of differential expression for the ten divergently dysregulated genes that are dysregulated in the greatest number of cell types. FC - fold change.

Neuronal cells showed higher transcriptional diversity than glial cell types, and excitatory neuronal populations exhibited strong layer-specificity. Superficial-layer excitatory neurons (layers II/III) are marked with *CUX2* and *CBLN2*; layer IV/V neurons with *RORB*, and *RXFP1*; and deep layer neurons with *TLE4*, *SEMA3E*, and *HTR2C* gene expression. Within cortical layers V and VI we identified three distinct populations of excitatory neurons: cortico-fugal projection neurons (Ex-L5/6) expressing *FEZF2*, and two distinct populations of cortico-cortical projection neurons expressing *NTNG2* (Ex-L56_CC_NTNG2) and *SEMA3A* (Ex-L6_CC_SEMA3A) (**Fig. 1b**). GABAergic inhibitory neurons are organized in major subtypes, including calcium-binding protein parvalbumin (*PV*) expressing neurons, neuropeptide somatostatin (*SST*) neurons, and the ionotropic serotonin receptor 5HT3a (5HT3aR) neurons. Within these groups, we detected two PV-expressing subtypes of inhibitory neurons (Basket and Chandelier), two 5HT3aR-expressing subtypes (VIP+ and Reelin+), and two subpopulations of the recently-described Rosehip interneurons (34), and we validated this heterogeneity within Rosehip interneurons using in situ hybridization (**Extended Data Figure 3**). The relative position of the cells within the cell similarity network is consistent with the developmental origin of cardinal interneuron subtypes (medial versus caudal ganglionic eminence), suggesting a developmental basis for shared transcriptional signatures (35).

**Figure 3.**
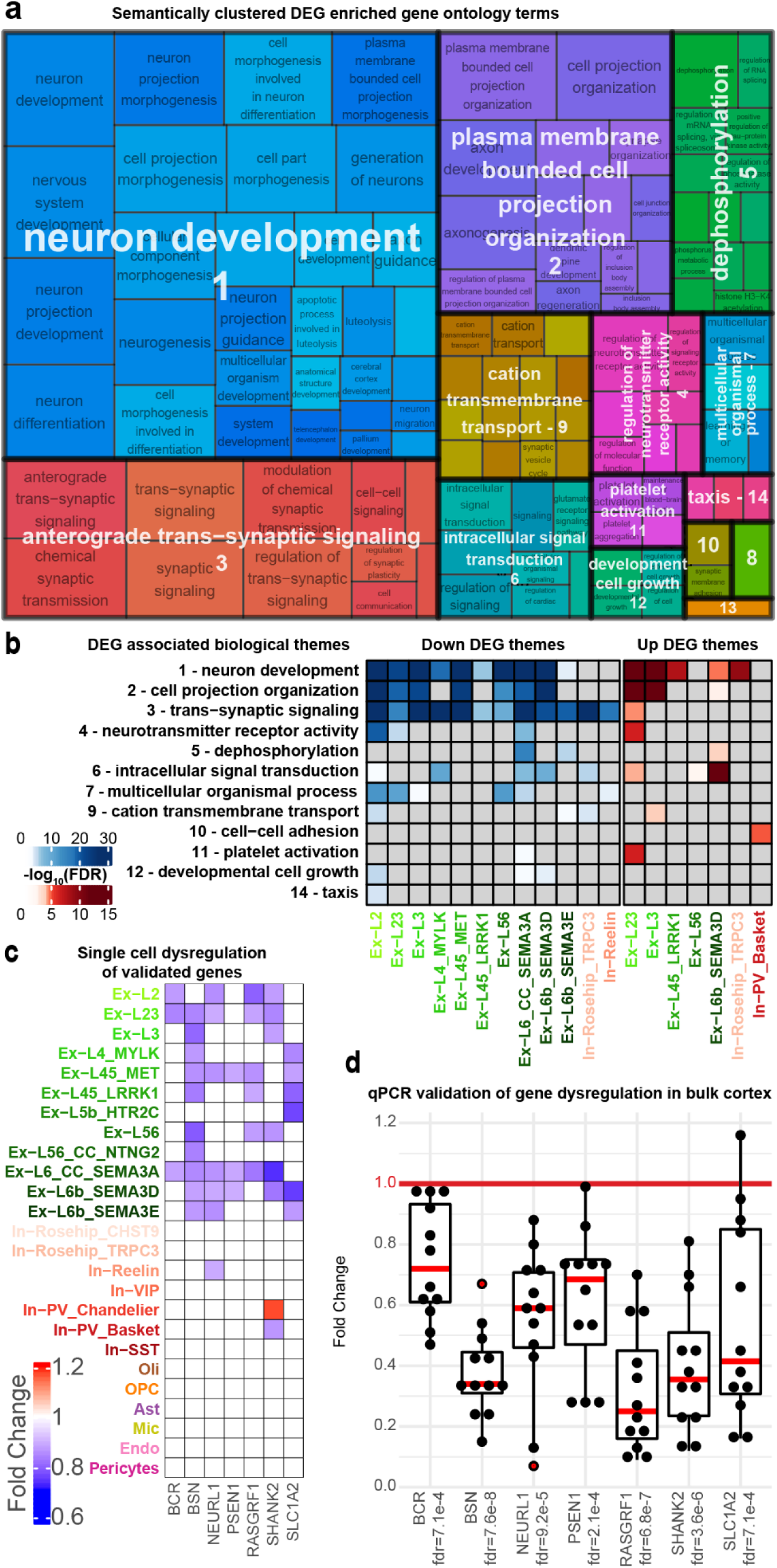
Associations of DEGs with biological pathways and qPCR validation. **a.** Biological pathway enrichment by down and upregulated cell type-specific DEGs determined by unbiased gene set enrichment analysis of all GO terms followed by semantic clustering of 119 enriched GO terms revealed 14 distinct biological themes named for the most significantly enriched GO term. Numbers indicate ordering of themes by the significance of the most significant term they contain. 8 - “chaperone-mediated protein folding”; 10 - “cell-cell adhesion via plasma-membrane adhesion molecules”; 13 - “behavior”. **b.** Aggregate enrichment of GO terms within each biological theme across cell types by down (blue) and upregulated (red) DEGs. Themes are ordered by the significance of their most enriched GO term. Two themes are not included in panel b that were implicated by only one (“chaperone-mediated protein folding” associated with upregulated DEGs in Ex-L6_CC_SEMA3A) any by two terms (“behavior” associated with downregulated DEGs in Ex-L56 and In-Rosehip_TRPC3). Cell types with no FDR significantly enriched themes are not depicted. **c.** Cell type-specific differential expression profiles for seven genes within the snRNA-seq meta-analysis results were selected for RT-qPCR validation. Only significant expression changes (FDR < 0.05 and abs(log_2_(fold change)) > 0.1) are depicted. **d.** qPCR validation of decreased expression of seven genes represented within multiple significantly enriched GO terms within the synaptic signaling theme, performed in RNA extracted from whole cortex tissue samples of 12 schizophrenia and 11 control subjects within the McL cohort. Black dots indicate fold change of expression of each target gene in one schizophrenia subject relative to average expression across all control subjects normalized to 1, computed with the ΔΔct method and beta-2 microglobulin (*B2M*) used as the reference gene. Red dots indicate outlier values.

While the literature is inconsistent, prior histologic studies have suggested loss of cells within inhibitory neuronal populations in schizophrenia (36), with much attention focused on parvalbumin-expressing interneurons (37). Prior studies were performed using histology-based cell-counting techniques, and current thinking interprets the detected neuronal loss as due to technical limitations of these studies, where marker expression fell below the level of histological detection without loss of cells (38). The current data generated by the unbiased sampling of cells across all cortical layers support this conclusion, as we detected no change in the representation of any cell type, including all subtypes of inhibitory neurons, between schizophrenia and control subjects (**Extended Data Fig. 1g**).

### Cell-type-specific expression changes in schizophrenia

To identify transcriptional alterations associated with schizophrenia, we systematically performed differential expression analysis for each cell type in each cohort separately, and subsequently performed a meta-analysis to integrate the results (Methods). Meta-analysis identified a total of 6,634 differential expression events (FDR < 0.05 and |log_2_(fold change)| > 0.1, 2,455 unique DEGs) across all cell types, most of which (77%, n=5,141) are downregulated in schizophrenia (**Fig. 2a**, Supplementary Table 4). Changes occur predominantly in neuronal populations (94%, n=6,229) and mostly in excitatory neurons (77%, n=5,129). Overall, DEGs were reproducible across datasets (**Fig. 2b**), with 745 genes consistently detected as significantly up or downregulated in both cohorts independently. Cross-cohort DEG reproducibility varied across cell types and was highest in excitatory neuron subpopulations. DEGs identified by means of meta-analysis significantly overlapped with those identified in each cohort separately, supporting the data integration procedure (**Fig. 2b**, Supplementary Table 5), and we focused on meta-analysis results for downstream analyses. We report cohort-specific analyses, a joint meta-analysis of differential expression across 25 cell types, and a highly reproducible set of 287 DEGs with consistent expression changes in all three analyses (Supplementary Table 6).

Comparative analysis with schizophrenia DEGs identified in high-quality bulk cortex RNA-seq data from the PsychENCODE consortium (42) (559 schizophrenia vs. 936 control samples finding 4,821 DEGs with FDR < 0.05) validated the DEGs that we identified in our snRNA-seq data, with a high concordance of expression changes, especially with those observed in neurons. Enrichment analysis revealed that bulk data primarily capture expression changes observed at the single-cell level in excitatory neurons, astrocytes, and oligodendrocytes (**Fig. 2c**). Notably, changes in other cell types, including inhibitory neurons, are less well captured, highlighting the relevance of single-cell assessments.

Most upregulated DEGs occurred in excitatory neurons of the superficial cortical layers, while the most downregulation events occurred in deep layer excitatory neurons, consistent with prior mapping of bulk schizophrenia DEGs to cortical layers based on spatial transcriptomics (43) (**Fig. 2a**). Nearly half of DEGs are perturbed in a single cell type (47%, n=1,153), indicating high cell-type specificity of expression changes. Schizophrenia cell type-specific DEGs do not overlap significantly with genes responsive to chronic antipsychotic exposure in non-human primates (44) (**Extended Data Fig. 4**), suggesting that identified expression changes are not driven by medication exposure.

**Figure 4.**
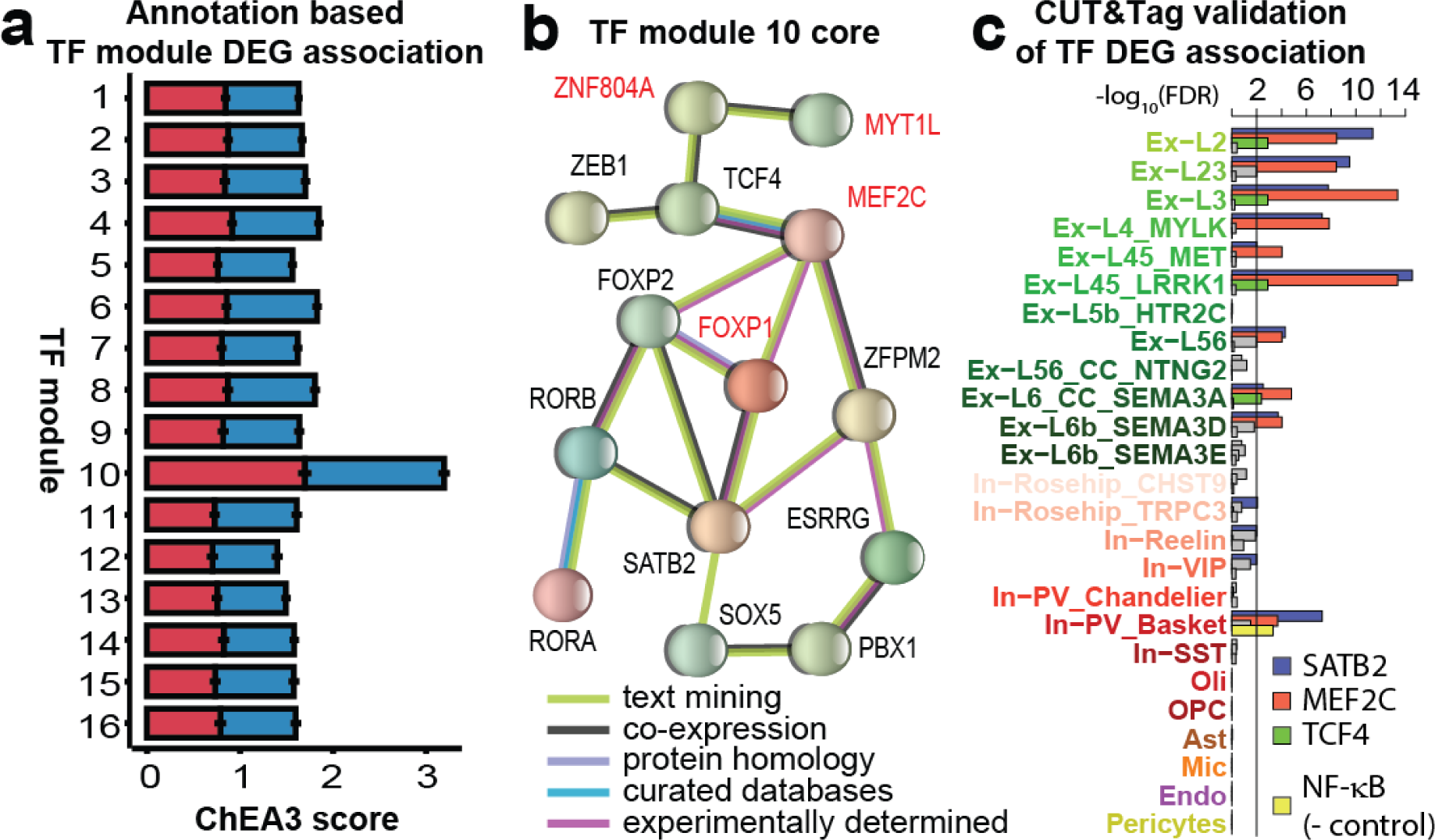
Schizophrenia DEGs implicate a coherently expressed TF module. **a.** TF co-expression across all individuals and all neuronal populations identified 16 coherently expressed modules ordered by size (module 1 is the largest). ChEA3 (62) analysis aggregated across all neuronal cell types and all TFs within each module reveals the greatest relatedness of module 10 to schizophrenia DEGs. Red bars depict the ChEA3 score for upregulated DEGs, blue bars depict the ChEA3 score for downregulated DEGs. **b.** TFs within module 10 form a highly connected protein-protein association network based on multiple lines of evidence curated within the STRING database (51). TFs named in red are fine-mapping prioritized schizophrenia risk genes reported by the Psychiatric Genomics Consortium (15). **c.** Genomic loci bound by MEF2C, SATB2, or TCF4 as observed in neuronal nuclei isolated from postmortem human prefrontal cortex with fluorescence-activated nuclear sorting are significantly associated with schizophrenia DEGs across multiple neuronal and no non-neuronal populations.

Among genes dysregulated in multiple cell types, 114 genes showed divergent directionality of changes across cell populations. The most broadly divergently dysregulated genes include *NRXN3*, *PFKFB3*, *RASGRF2*, *SHANK2*, and *DLG5*, all previously associated with schizophrenia by function or genetic data (42, 45–49). Our single-cell resolution data indicate that alterations in these genes tend to occur in a single direction within and divergently across cell classes (excitatory, inhibitory, and glial), suggesting that cell type-specific regulatory complexity may be relevant for gene expression modulation (**Fig. 2d**, **Extended Data Fig. 5**). Overrepresentation of genes involved in synaptic structure and function further suggests the biological relevance of divergent dysregulation across neuronal classes.

**Figure 5.**
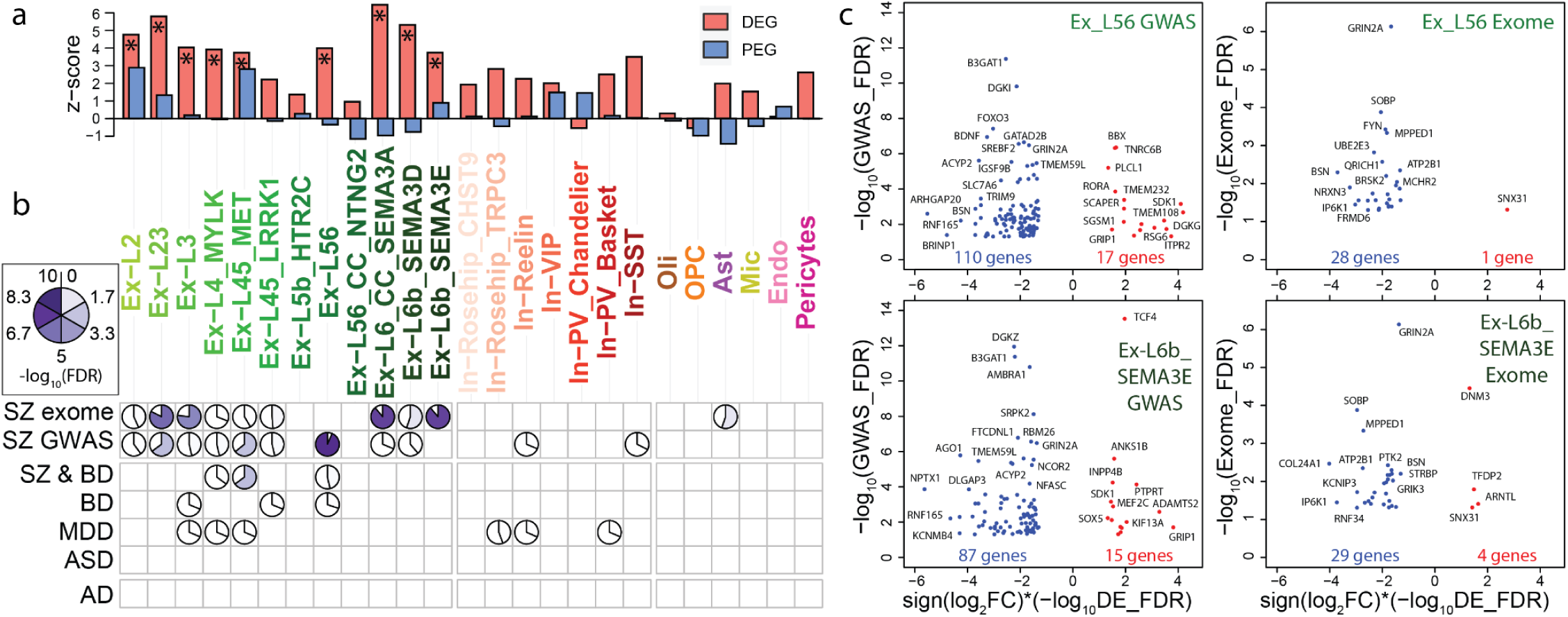
Association of differentially expressed genes with genetic risk variants across cell types and disorders. **a.** Enrichment of GWAS signal with individual cell-type-specific schizophrenia DEGs (red) and equal numbers of genes preferentially expressed in each cell type (PEG - blue) assessed by permutation test against randomly selected genes. Asterisks indicate Bonferroni significant difference (p>0.01) between the DEG and random gene sets. PEG and random gene sets are not significantly different in any cell type. **b.** Significance of association genome wide between cell-type-specific schizophrenia differential expression and genomic variants implicated by GWAS in six distinct neuropsychiatric disorders computed with H-MAGMA (72) and by exome sequencing for schizophrenia (71) (SZ exome) computed by GSEA. **c.** Visualization of genes most strongly associated (FDR < 0.05) with common schizophrenia risk variants (GWAS) computed with H-MAGMA (y-axis, left) or rare schizophrenia risk variants (exome sequencing, y-axis, right) with significant expression changes in schizophrenia (x-axis) within deep layer excitatory neuronal populations showing the strongest associations in panel b.

### Differentially expressed genes impact neurodevelopment and synapse related pathways

To determine the biological pathways most likely impacted by these cell type-specific dysregulated gene sets, we performed gene ontology analysis for the 50 DEG sets (up and downregulated genes considered separately for each of 25 cell types) and semantically clustered the significantly enriched terms into biological themes to aid interpretation. Observed expression changes converged to 14 biological themes summarizing functionally related molecular pathways, with neurodevelopmental and synaptic processes highly overrepresented within schizophrenia DEG associations (**Fig. 3a, b**, Supplementary Table 7). Biological themes related to neurodevelopmental processes were associated with DEG sets within excitatory and inhibitory neuronal populations with the terms “neuron development” and “plasma membrane-bound cell projection organization” most significantly enriched.

Within synapse-related processes the theme “anterograde trans-synaptic signaling” was the most broadly perturbed, associated with downregulated DEG sets in 12 distinct neuronal populations and with upregulated DEG sets within superficial layer excitatory neurons Ex-L2 and Ex-L23. The theme “regulation of neurotransmitter receptor activity” was dysregulated within more select populations of excitatory neurons, and included more specific terms related to glutamatergic signaling through regulation of AMPA and NMDA receptors. We validated the downregulation of seven genes broadly represented across multiple pathways within these synaptic relevant themes (*BCR*, *BSN*, *NEURL1*, *PSEN1*, *RASGRF1*, *SHANK2*, *SLC1A2*) with quantitative PCR with reverse transcription (qPCR - **Fig.3c**, **d**). All seven genes were found to be significantly downregulated in schizophrenia when comparing RNA extracted from whole cortex postmortem BA10 tissue from 12 schizophrenia versus 11 control subjects (McLean cohort).

Across all cellular populations, Ex-L23 upregulated DEGs significantly enriched the greatest number of themes (6 of 14), consistent with having the largest number of upregulated DEGs, while downregulated DEGs within Ex-L2 and Ex-L6_SEMA3A enriched the greatest number of themes (9 of 14), despite Ex-L2 having less than half the number of downregulated DEGs as some deep layer excitatory populations. Inhibitory neurons showed more modest perturbation of these themes, with synaptic signaling enriched by downregulated DEGs in In-Rosehip_TRPC3 and In-Reelin, and neurodevelopment relevant terms enriched by upregulated DEGs in In-Rosehip_TRPC3. Altogether, our data suggest that schizophrenia DEGs do converge on functionally related molecular pathways, with the strongest evidence implicating neurodevelopmental, synaptic, and intracellular signaling relevant processes.

### Neuronal DEGs are targets of schizophrenia GWAS risk transcription factors

Coordinated expression changes are often driven by common upstream transcriptional regulators (50), and we investigated whether observed cell-type specific dysregulation was associated with specific trans-acting factors. We grouped regulators with similar expression profiles by co-expression analysis (Supplementary Table 8) and performed TF target analysis to statistically assess the overrepresentation of target genes within schizophrenia DEGs, and identified a single TF module highly associated with neuronal DEGs (TF module 10, **Fig. 4a**). This module includes 24 TFs coherently expressed and supported by protein-protein interactions documented in the literature, with 14 of these TFs linked in a highly-connected interaction network supported by multiple lines of evidence (51) (**Fig. 4b**).

TFs within this module are involved in neurodevelopment and overlap (7 of 24) with TFs encoded by schizophrenia GWAS risk genes (15). *MYT1L* encodes a nervous system-specific TF that represses non-neuronal genes during neurodevelopment (15). *MYT1L* is linked to schizophrenia through GWAS fine mapping and by gain of function mutations, and to autism spectrum disorders through loss of function (52). *SATB2* encodes a transcription factor active in chromatin remodeling and large-scale genome architecture that plays a role in excitatory neuron subtype specification (53) and regulates genes linked to schizophrenia, educational attainment, autism, and intellectual disability (54)*. TCF4* is a broadly expressed helix-loop-helix transcription factor involved in nervous system development linked to the severe neurodevelopmental disorder Pitt Hopkins Syndrome (55) that has been previously predicted as a “master regulator” of schizophrenia gene network perturbations (56). *MEF2C* binds a DNA motif enriched near schizophrenia genomic risk loci, enhances cognition when overexpressed in mice (57), and is associated with excitatory/inhibitory balance in the developing brain (58). *SOX5* is implicated in fate determination and regulation of corticofugal projection neuron development, with loss-of-function perturbations leading to disrupted neuronal proportions and emergence timing (59). *ZNF804A* is a schizophrenia fine-mapping prioritized GWAS gene (15) involved in neurodevelopment, synapse development, and dendritic morphology (60).

To validate predicted associations between prioritized TFs and observed schizophrenia DEGs, we experimentally tested whether active cis-regulatory elements that are targeted by these TFs show preferential association with schizophrenia DEGs. We performed Cleavage Under Targets and Tagmentation (CUT&Tag (61)) assays in four control individuals within the McLean cohort to map the genome-wide binding of MEF2C, SATB2, and TCF4, and further included NFKB as a negative control not expected to show strong association with schizophrenia DEGs. These assays were performed in neuronal nuclei isolated from postmortem prefrontal cortex tissue samples using fluorescence-activated nuclear sorting. We defined reproducibly bound regions as those bound in at least 50% of replicates and annotated them with the nearest gene. Supporting the specificity of this analysis, genes targeted by MEF2C, SATB2, and TCF4 preferentially enrich biological pathways relevant to neuronal function (**Extended Data Fig. 6**). We found a high degree of overlap between identified target genes and schizophrenia DEGs across a wide range of excitatory neurons, with less but still significant overlap in multiple inhibitory populations (**Fig. 4c**). MEF2C was strongly associated with schizophrenia DEGs across nine excitatory populations and In-PV_Basket, and SATB2 was strongly associated with schizophrenia DEGs within the same excitatory populations and four inhibitory cell types.

**Figure 6.**
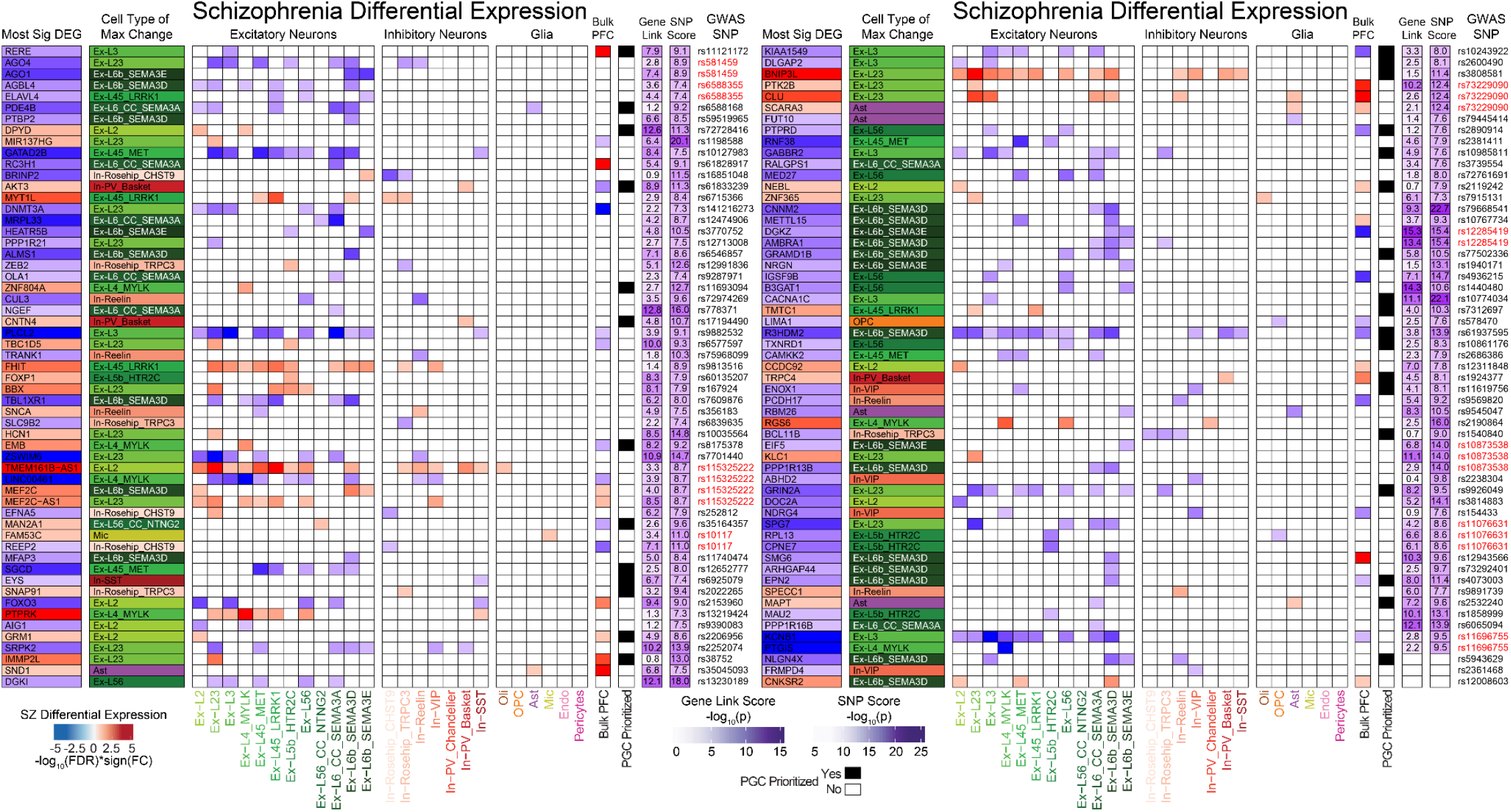
Cell type-specific differential expression of high confidence schizophrenia risk genes. Shown are the 114 genes within the Psychiatric Genomics Consortium’s broad fine-mapped set (15) that were differentially expressed in at least one cell type, depicting the direction of change (red - upregulated; blue - downregulated), the cell type in which the most significant DE event occurs, and a heatmap depicting significant (FDR < 0.05 and abs(log_2_(fold change)) > 0.1) dysregulation across cell types and in prior bulk PFC assessment (42). Purple columns indicate the significance of the association of each gene with schizophrenia common risk loci computed with H-MAGMA (Gene Link Score), and the significance of the index SNP linked to each gene by the PGC3’s statistical fine mapping (SNP Score). PGC prioritized indicates if a gene is present within the Psychiatric Genomics Consortium’s set of 120 prioritized schizophrenia risk genes. Index SNPs in red are linked to multiple DEGs. Figure is split only for space considerations.

TCF4 was more modestly associated with schizophrenia DEGs within four excitatory populations only. None of the tested TFs showed a strong association with DEGs in any non-neuronal cell-type. As expected, NFKB was less associated with schizophrenia DEGs in all cell types, although it was significantly associated with DEGs in In-PV_Basket. Overall, this experimental data supports the evidence of a regulatory association between members of TF module 10 and cell type-specific schizophrenia dysregulated genes.

### Schizophrenia DEGs are associated with common and rare genetic risk variants

Seeking insights into the association between transcriptional alterations and genetic liability to schizophrenia, we investigated the relationship between transcriptionally dysregulated genes and genetic risk loci. In this context, two questions are of relevance. First, genes that are dysregulated in schizophrenia may also be associated with genetic variation, suggesting a link between genetic liability and measurable cell type-specific molecular alterations. Second, genes that are preferentially expressed by specific cell types may, as a class, provide schizophrenia risk, suggesting a functional role of those cells in mediating genetic risk. We found strong evidence supporting the first scenario in most excitatory neurons, and only modest evidence for the second in superficial-layer neurons (**Fig 5a**, **Extended Data Fig. 7**, Supplementary Table 9). Gene-level genetic risk scores computed from GWAS summary statistics (see Methods) were unexpectedly high for DEGs from 9 out of 12 excitatory subpopulations (Bonferroni-corrected p-value<0.01, resampling test), with Ex-L6_CC_SEMA3A neurons showing the strongest deviation from random expectation. Notably, we did not observe similar associations for preferentially expressed genes (PEGs), which showed only modest evidence for higher than expected GWAS risk scores (nominal p-value<0.01) in excitatory subpopulations of layers II, IV, and V. Consistent with these observations, GWAS scores of schizophrenia DEGs overall tend to be higher than those of genes preferentially expressed in the same cell type (nominal p-value<0.05 in most cell types, Bonferroni-corrected p-value<0.01 in deep-layer excitatory neurons), and only a single cell type, In-PV_Chandelier, displayed higher values for preferentially expressed genes.

Past efforts have identified potential cell types impacted by disease-associated genetic variants by assigning schizophrenia risk genes to cell types that preferentially express them in neurotypical brains (15, 63). Our data suggest this approach has merit, as preferentially expressed genes are more associated with genetic risk than are randomly selected genes in relevant neuronal subtypes. With the caveat that genetic risk loci may influence gene function through additional mechanisms beyond transcriptional dysregulation, and that transcriptional alterations observed in postmortem cortical samples of schizophrenia subjects might stem from several direct and indirect mechanisms, our data complement previous findings supporting a degree of convergence in genetic and transcriptional perturbations in schizophrenia that is largely cell-type specific.

To further investigate the association between transcriptional alterations and genetic liability to schizophrenia genome-wide, we used Hi-C-coupled MAGMA (H-MAGMA) (64) to first assign non-coding SNPs to their cognate genes based on evidence of long-range interactions in the adult dorsolateral prefrontal cortex (65) and then test the statistical association between aggregate gene-level genetic risk from common variants and cell type-specific expression alterations. In addition to schizophrenia (15), we further considered risk loci for four psychiatric disorders that are known to share genetic risk factors (64): major depressive disorder (MDD) (66), bipolar disorder (BD) (67), autism spectrum disorder (ASD) (68), and attention-deficit/hyperactivity disorder (ADHD) (69). This approach allows us to distinguish schizophrenia from general psychiatric illness-related associations. As a point of contrast, we included Alzheimer’s disease (AD) (70), a neurodegenerative disorder not expected to be genetically related to schizophrenia. Furthermore, to extend associations beyond common, small effect variants, we also considered gene associations by overrepresentation of rare protein coding variants in schizophrenia subjects (SCHEMA) (71).

We found strong associations between schizophrenia DEGs and genes targeted by common or rare risk variants in multiple neuronal subpopulations, suggesting substantial causal effects in observed transcriptional alterations (**Fig. 5b**). Across excitatory neuronal subpopulations, the observed layer-specificity of these associations was again consistent with the enrichment of schizophrenia genetic variants proximal to genes preferentially expressed in layer II and layer V (43). While common and rare risk variants were associated with a highly overlapping set of excitatory neuronal populations, separations did occur with Ex-L56 DEGs being the most significantly associated with common variants (GWAS), but not significantly associated with rare protein coding mutations (SCHEMA). Conversely, Ex-L6b_SEMA3E was the most significant association with rare variants but was not significantly enriched with common variants. As the gene sets and biological processes impacted by common and rare variants are thought to be highly overlapping (71), the divergence of excitatory neuronal populations showing expression changes in associated genes observed here was unexpected and potentially relevant to understanding how the outcomes of distinct categories of genetic risk factors are partitioned across the cortical cytoarchitecture. While these associations were predominantly observed within excitatory populations, there was significant association between GWAS variants and DEGs within two inhibitory populations, and the only significant association seen within non-neuronal populations was between astrocyte DEGs and rare protein-coding mutations, in agreement with prior evidence that these cell types play a less prominent role in schizophrenia pathologies mediated by genetic variation (63).

We found that some of the cell types whose transcriptional perturbations show a strong association with schizophrenia genetic risk variants are also associated with risk variants for bipolar and major depressive disorders, which is consistent with their previously-reported strong genetic relationships both at the level of genetic correlations and gene-level overlaps (72). Bipolar disorder-associated variants showed overlap with schizophrenia DEGs within excitatory populations Ex-L3, Ex-L45_LRRK1, and Ex-L56, while those for major depressive disorder overlap with excitatory populations across layers III, IV, and V, as well as three inhibitory neuronal populations. We found no significant associations between schizophrenia transcriptional perturbations and ASD risk (72, 73), consistent with the known lower correlation of genetic risk between these disorders (73) (with the caveat that the ASD GWAS study on which this analysis is based is less well powered than those used for other disorders). As expected, we found no overlap with genetic risk for the neurodegenerative disorder AD.

We next examined specific genes with strong evidence of both an association with genetic risk and differential expression within neuronal populations with convergent genetic and transcriptomic alterations (**Fig. 5c**, **Extended Data Fig. 8**). We observed consistently strong downregulation of *GATAD2B*, encoding a zinc finger transcription factor involved with transcriptional repression through deacetylation of histones in proximity to methylated DNA implicated in neurodevelopmental disorders (74). The related genes *DGKI* and *DGKZ* were also consistently downregulated, encoding diacylglycerol kinases active in multiple lipid signaling pathways regulating neuronal function (75). Within the same gene family, *DGKG* was upregulated across 11 excitatory populations and in OPC. *FXR1* was strongly upregulated in multiple populations, and encodes an RNA binding protein functionally related to fragile-x mental retardation protein (76). *HSPA1A* was also upregulated and encodes a heat shock protein linked to schizophrenia-relevant neurodevelopmental processes and associated with schizophrenia symptom severity (77). *TCF4*, discussed earlier, was strongly upregulated across multiple neuronal populations.

Shifting to genes associated with ultra-rare protein-coding variants in schizophrenia, we observed widespread downregulation of *GRIN2A* and *NRXN3*, both discussed above. Also broadly downregulated was *BSN*, encoding bassoon, aka presynaptic cytomatrix protein, active in structural regulation of the presynaptic active zone (78), and *SOBP*, encoding a neurodevelopment-related zinc finger transcription factor linked to intellectual disability (79). The most widely upregulated gene also implicated by exome sequencing in schizophrenia was *SNX31*, upregulated in 10 excitatory populations, encoding a sorting nexin involved in the regulation of protein trafficking. *DNM3* was also upregulated across multiple neuronal populations, and encodes dynamin 3, a force producing protein involved in the regulation of synaptic endocytosis (80) and dendritic spine morphology.

Focusing on genes linked to credible, common schizophrenia-associated variants, we observed strong evidence of differential expression within at least one cell type for 114 of the Psychiatric Genomics Consortium’s “broad fine-mapped set” of 628 schizophrenia risk genes (15), mapping transcriptional dysregulation of these genes across cell populations within the human prefrontal cortex (**Fig. 6**). Among these 114 genes, 110 were protein coding genes, while two were antisense transcripts (MEF2C-AS1 and TMEM161B-AS1), and two were long intergenic noncoding RNAs (LINC00461 and MIR137HG). For 87 of these genes the most significant schizophrenia differential expression event was observed in an excitatory neuronal population, while 20 were most robustly altered in inhibitory neurons, and 7 in non-neuronal populations. 113 of the 628 prioritized genes were dysregulated in a past assessment of bulk prefrontal cortex (42), with 36 genes dysregulated in both the bulk cortex and the current single cell study. The Psychiatric Genomics Consortium further refined their gene list using fine-mapping, summary-based Mendelian randomization, and eQTL colocalization analysis to prioritize 120 genes presumed to mediate schizophrenia risk. We observed differential expression of 31 of these genes in the present study.

Genes and SNPs exhibit a many-to-many relationship, and there were nine PGC3 index SNPs mapped to multiple dysregulated genes. Gene sets linked to a single index SNP showed a tendency to be dysregulated with the same directionality, although three SNPs were linked to both up and downregulated genes, and all of these genes were dysregulated in a single direction across cell-types. SNP rs115325222 was linked to upregulated TMEM161B_AS1, MEF2C, and MEF2C-AS1, as well as downregulated LINC00461. SNP rs10117 was linked to upregulated FAM53C and downregulated REEP2. SNP rs10873538 was linked to downregulated EIF5 and PP1R13B as well as upregulated KLC1. The Psychiatric Genomics Consortium analysis (15) identified 287 genomic regions linked to common risk variants for schizophrenia, and we observed between one and three schizophrenia DEGs within 88 of these regions. As has been demonstrated for ultra-rare protein-coding genomic variants (71), disease-associated transcriptomic changes may prove useful in the prioritization of genes within schizophrenia GWAS loci.

Illuminating the complexity of disease-associated gene expression changes across the cytoarchitecture of the human cortex, we observed five fine-mapped schizophrenia risk genes to be divergently dysregulated across cell types, including *BRINP2*, *EFNA5*, *MAPT*, *SNCA*, and *ZEB2* (**Fig. 6**). This observation demonstrates that even genes presumed to be strongly linked to disease-associated genetic elements are influenced by cell type-specific regulatory mechanisms. Among these divergently dysregulated fine mapped genes, *BRINP2* encodes a neuron-specific developmentally regulated cell cycle regulatory protein (81), and was upregulated in Ex-L6b_SEMA3E while being downregulated in both observed populations of Rosehip neurons. *EFNA5* encodes Ephrin-A5, a cell surface protein ligand of Eph receptor tyrosine kinases involved in cell-cell interactions mediating axon guidance and neuronal plasticity in adulthood (82), and was downregulated in In-Rosehip_CHST9 and upregulated in Ex-L23. *MAPT* encodes microtubule-associated protein tau important in maintenance of neuronal polarity and implicated in multiple neurodegenerative disorders (83) which was downregulated in Ex-L6_CC_SEMA3A and upregulated in astrocytes. *SNCA* encodes alpha-synuclein, a regulator of synaptic vesicle trafficking and neurotransmitter release (84), and was downregulated in Ex-L45_MET and upregulated in In-Reelin. *ZEB2* encodes a zinc finger transcriptional repressor involved in neurodevelopment and dopaminergic axon growth and target innervation (85), and was downregulated in In-Rosehip_TRPC3 and upregulated in Ex-L5b_HTR2C.

In addition to GWAS-identified common schizophrenia risk variants, we also assessed genes strongly associated with rare protein-coding variants in schizophrenia for cell type-specific differential expression (71). Among the 32 genes associated at FDR < 0.05 we found nine to be differentially expressed, primarily in excitatory neuronal populations, with only one gene dysregulated exclusively in a non-excitatory neuron population (**Extended Data Fig. 9**). Of the ten genes with exome-wide significant association, four were differentially expressed, exclusively through downregulation in schizophrenia. *GRIN2A* was downregulated in seven excitatory neuronal populations and in *TRPC3* expressing Rosehip interneurons, is also a fine-mapping prioritized schizophrenia risk gene, and encodes a subunit of the NMDA receptor of central importance to synaptic plasticity (86). *GRIA3* was downregulated in Ex-L6b_SEMA3E and in OPC, and encodes a subunit of the AMPA receptor involved in regulating synaptic strength through multiple mechanisms (87). *CACNA1G* was downregulated in Ex-L56_CC_NTNG2 neurons and encodes a low-voltage-activated or T-type calcium channel implicated in neurotransmitter release and in action potential bursting (88). *HERC1* was downregulated in Ex-L45_LRRK1 neurons and encodes a guanine exchange factor for ARF1 and Rab proteins active in membrane trafficking mechanisms that has been linked to intellectual disability (89).

### Transcriptional pathology recovers heterogeneous schizophrenia subgroups

Schizophrenia is a highly heterogeneous disorder, with classical descriptions of multiple clinical subtypes and evidence of diverse etiological factors. To investigate whether such heterogeneity manifests at the transcriptomic level across cell-types and subjects within the present cohorts, we developed a transcriptional pathology score (TPS). TPS assesses the degree of consistency between the genome-wide gene expression of each individual (relative to the average across all individuals) and the expression changes observed between schizophrenia and control subjects, therefore allowing the estimation of discrepancies between expression profiles and diagnostic categories, and complementing the binary case versus control diagnosis with a continuous measure of transcriptomic change. A high TPS value indicates that the relative expression profile of an individual is globally consistent with the expression changes expected in schizophrenia compared to control subjects. TPS were calculated for each cell type separately, and each individual was assigned an aggregate TPS averaged across neuronal cell types (Methods). By ranking all subjects by their aggregate TPS we discovered four groups of individuals: schizophrenia subjects that appear transcriptionally consistent with schizophrenia (SZ), schizophrenia subjects that appear transcriptionally more similar to control subjects (SZ_CON-like), control subjects that appear transcriptionally consistent with control (CON), and control subjects that appear transcriptionally more similar to the schizophrenia group (CON_SZ-like) (**Fig. 7a**, **b**). This analysis demonstrates that clinical diagnosis and underlying transcriptional patterns are not always consistent, suggesting structured heterogeneity within diagnostic groups at the transcriptional level. In particular, a subset of clinically diagnosed schizophrenia subjects do not present the expected transcriptional alterations observed across the whole cohort, consistent with prior studies of gene expression in bulk cortex (90).

**Figure 7.**
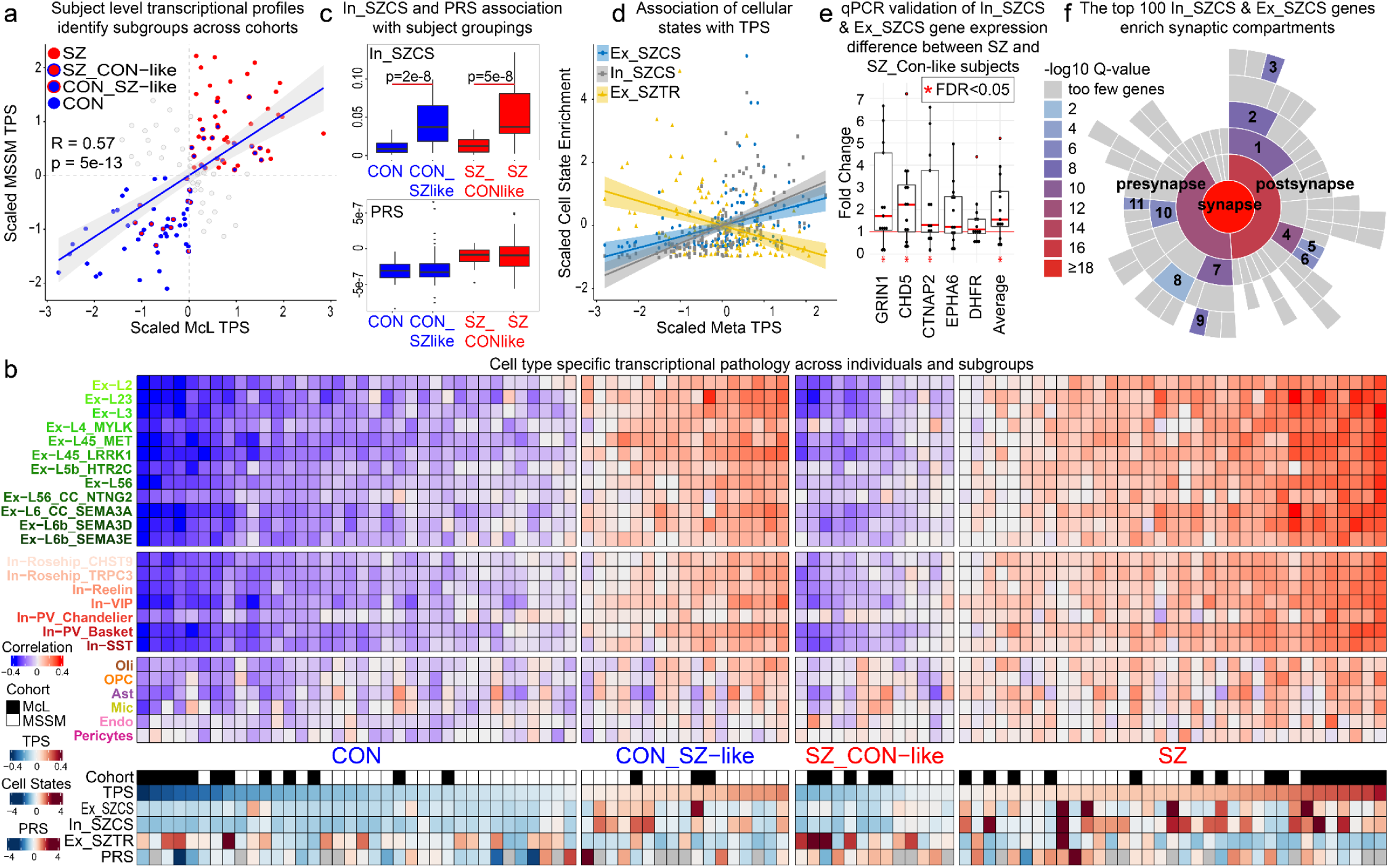
Heterogeneity of transcriptional changes across individual subjects. **a.** Individual subjects plotted by their degree of schizophrenia-associated transcriptional pathology computed within the space of the schizophrenia-associated transcriptional changes observed in the McL cohort (x-axis) or the MSSM cohort (y-axis). Values are scaled, with larger values indicating a greater association with schizophrenia. Four subgroups of subjects are identified: schizophrenia subjects with a transcriptional profile consistent with schizophrenia within both cohorts (red points in the upper right quadrant), control subjects with a transcriptional profile consistent with control within both cohorts (blue points in the lower left quadrant), schizophrenia subjects with a transcriptional profile consistent with control within both cohorts (red points outlined in blue in the lower left quadrant), and control subjects with a transcriptional profile consistent with schizophrenia within both cohorts (blue points outlined in red in the upper right quadrant). Off-diagonal subjects (gray) are inconsistent between analyses (appearing transcriptionally similar to control in one and schizophrenia in the other) and were not considered further in this analysis. **b.** Cell type-specific schizophrenia-associated transcriptional pathology within subjects clustered into the four groups identified in panel a. Values are scaled with darker red indicating a value more characteristic of the schizophrenia group, and darker blue indicating a value more characteristic of the control group. Correlation - cell type-specific correlation between each subject’s transcriptional signature and the transcriptional signature characteristic of the SZ group; TPS - Transcriptional Pathology Score averaged across all cell types for each individual; Ex_SZCS, In_SZCS, Ex_SZTR - scaled expression of each transcriptional signature within each subject; PRS - schizophrenia Polygenic Risk Score. Subjects with inconsistent classification in panel a were omitted from panel b. **c.** Boxplots depict the enrichment of the In_SZCS transcriptional signature (top) within all subjects (McL and MSSM combined) across the four groups identified in panel a, showing high association with subject grouping. Subject PRS (bottom) does not show this association. **d.** Scaled scatterplot demonstrating the strong correlation between cell-state transcriptional signatures and schizophrenia transcriptional pathology across subjects, with In_SZCS and Ex_SZCS (R=0.56, p=4.6e-13, and R=0.35, p=1.9e-5, respectively) showing positive and Ex_SZTR showing inverse association with TPS (R=-0.39, p=1.8e-6). **e.** qPCR validation of increased expression of five top In_SZCS associated genes in SZ as compared to SZ_CON-like subjects, performed in RNA extracted from whole cortex tissue samples of 13 SZ and five SZ_CON-like subjects within the McL cohort. Black dots indicate fold change of expression of each target gene in one SZ subject relative to average expression across all SZ_CON-like subjects normalized to 1, computed with the ΔΔct method and beta-2 microglobulin (*B2M*) used as the reference gene. Red dots indicate outlier values. **f.** The top 100 genes characterizing both the Ex_SZCS and In_SZCS transcriptional signatures encode proteins localized to the synaptic compartment as annotated within the SynGO database(94). 1 - postsynaptic specialization; 2 - postsynaptic density; 3 - integral component of postsynaptic density membrane; 4 - postsynaptic membrane; 5 - extrinsic component of postsynaptic membrane; 6 - integral component of postsynaptic membrane; 7 - presynaptic active zone; 8 - synaptic vesicle membrane; 9 - integral component of synaptic active zone membrane; 10 - presynaptic membrane; 11 - integral component of presynaptic membrane.

TPS is computed based on reference case-control expression changes. To test whether similar patterns of inconsistency between diagnosis and transcriptional pathology occur in both cohorts, we performed the following computational validation experiment. We classified subjects into SZ, SZ_CON-like, CON, or CON_SZ-like based on TPS calculated using schizophrenia-associated expression changes observed only within the McL cohort, only within the MSSM cohort, or jointly in the cross-cohort meta-analysis, and then quantified group assignment consistency across these classifications. In all cases we found significant overlap between subjects of each class (**Extended Data Fig. 10**), indicating patterns of inconsistency between diagnosis and transcriptional pathology across individuals are reproducible in both cohorts. Subjects whose classification depended on the source of schizophrenia expression differences had TPS of small magnitude, suggesting discrepancy in classification stems from a low assignment confidence and not lack of reproducibility (off-diagonal quadrants in **Fig. 7a**).

To test whether discovered diagnostic subgroups could be explained by differences in underlying genetic risk, we next investigated the relationship between subject grouping and genetic background. We computed polygenic risk scores (PRS) for the subset of genotyped subjects of European ancestry (n=99 subjects, 45 McL and 54 MSSM) and found a significant correlation between TPS and PRS (Pearson correlation = 0.26, p-value = 9.4 x 10^-3^) and significantly different scores of each in schizophrenia versus control subjects (p-value < 1 x 10^-5^, **Extended Data Fig. 11**), supporting a relationship between transcriptional pathology as defined herein and genetic risk. However, PRS was not different between SZ and SZ_CON-like, or between CON and CON_SZ-like groups (**Fig. 7c**), indicating that these subject groupings are not explained by known schizophrenia common variant genetic risk factors.

To better understand the gene signatures underlying subject grouping, we applied a matrix decomposition approach to simultaneously uncover hidden patterns of variability across all subjects and cells. Unlike TPS, this unbiased approach does not rely on prior knowledge of subject diagnosis, but rather learns from the data transcriptional patterns that might recapitulate the observed subject-level heterogeneity of schizophrenia associated transcriptional changes. The majority of transcriptional signatures identified corresponded to individual cell types, while four signatures were robustly expressed by cells within multiple neuronal populations, representing transcriptionally defined cellular states (**Extended Data Fig. 12**). One of these cellular states was marked by high expression of the *NRGN* gene, and has been observed in prior snRNA-seq studies of human brain (30). The remaining three cellular states, two marking excitatory and one marking inhibitory neuronal populations, were, to our knowledge, novel to the present analysis, and displayed contrasting associations with schizophrenia transcriptional changes.

Two of the novel cellular states displayed a positive association with TPS, with one marking primarily excitatory neuronal populations (Ex_SZCS - excitatory schizophrenia cell state, R=0.35, p=1.9e-5) and the other inhibitory neuronal populations (In_SZCS - inhibitory schizophrenia cell state, R=0.56, p=4.6e-13), while the final cellular state (Ex_SZTR - excitatory schizophrenia transcriptional reversal) was negatively associated with TPS (R=-0.39, p=1.8e-6). These associations were robust within the assembled cohort (**Fig. 7d**) and reproducible in each cohort independently (**Extended Data Fig. 13**). While they marked distinct classes of neurons, In_SZCS and Ex_SZCS were highly correlated with respect to both the genes characterizing these transcriptional states (R=0.6, p<2.2e-16) and the subjects in which they were most prominently expressed (R=0.7, p<2.2e-16) (**Extended Data Fig. 14**). Both In_SZCS and Ex_SZCS were associated with subject grouping, with low average value in CON and in SZ_Control-like subjects, and high average value in SZ and in CON_SZ-like subjects, with In_SZCS showing the strongest evidence of association. Ex_SZTR showed the opposite pattern, with high average value in SZ_CON-like and CON subjects (**Fig. 7c**, **Extended Data Fig. 15**). Signatures of all three cellular states were correlated with TPS, but not with PRS (In_SZCS R=4.3e-2, p=6.6e-1, Ex_SZCS R=3.6e-2, p=7.2e-1; Ex_SZTR R=-1.4e-1, p=1.5e-1), suggesting an association with aspects of schizophrenia separate from known common genetic risk variants. Gene sets characterizing these three transcriptional states (Supplementary Table 10) were enriched for schizophrenia DEGs across multiple neuronal populations, with Ex_SZCS the most strongly enriched for up and downregulated DEGs within excitatory neurons, In_SZCS more strongly associated with DEGs in inhibitory neurons than the other cell states, and Ex-SZTR marked only by downregulated DEGs across multiple excitatory neuronal populations (**Extended Data Fig. 16**).

Considering the similarities between Ex_SZCS and In_SZCS, we merged the sets of genes characterizing these transcriptional signatures for further investigation by first z-score normalizing each gene’s specificity score, and then averaging the normalized scores across signatures. We found the gene most strongly associated with Ex_SZCS, In_SZCS, and both combined to be *DHFR*, encoding dihydrofolate reductase, a key enzyme in one-carbon metabolism (91). Top genes characterizing the combined transcriptional signature also included *GRIN1*, encoding subunit 1 of the NMDA receptor; *EPHA6*, encoding a receptor tyrosine kinase implicated in axon guidance; *CHD5*, encoding a neuron specific ATP dependent chromatin remodeling enzyme (92) with evidence of protein coding mutations linked to schizophrenia risk (71); and *CNTNAP2*, encoding a neurexin protein necessary for clustering of potassium channels at nodes of Ranvier in myelinated axons previously implicated in schizophrenia and other psychiatric diseases (93). Expression differences for these top genes between subgroups of schizophrenia subjects were validated with qPCR, with all five tested genes showing increased expression in SZ as compared to SZ_CON-like subjects, as expected, and strong evidence of increased expression for three out of the five tested genes as well as for the average fold change of all five tested genes (**Fig. 7e**). More broadly, annotation of the top 100 merged cell state genes within the SynGO database (94) found strong evidence of overrepresentation of genes encoding proteins localized to both the presynaptic (GSEA p = 2.4e-13) and postsynaptic (GSEA p = 8.3 e-17) compartments (Fig. 7f, **Extended Data Fig. 17**). These results suggest that among neuronal molecular processes associated with schizophrenia, expression of genes involved in one-carbon metabolism and neuronal depolarization (e.g. NMDA receptors, voltage-gated potassium channels), and enriched at the synapse, may indicate distinct molecular pathologies in subsets of individuals assigned a clinical diagnosis of schizophrenia.

## Discussion

Here we present a robust and reproducible single-cell transcriptomic case-control dissection of schizophrenia across two datasets generated independently with similar methodologies from distinct donor cohorts. We produced a high-resolution dataset of 468,727 single-cell transcriptomes sampled from 140 individuals and annotated 27 neuronal and glial cell types, which we used to investigate cell-type-specific schizophrenia-dysregulated genes, pathways, and regulators. While we observed significant changes in gene expression within all detected cell types, the majority of observed changes occurred in neuronal populations, with more than three quarters of all observed changes occurring in excitatory neurons. Consistent with prior work, we observed decreased gene expression to occur preferentially within deep layers of the cortex, while increased gene expression favored superficial populations. DEGs were highly cell-type-specific including multiple genes significantly and divergently dysregulated -- i.e., upregulated in some and downregulated in other cell populations. Half of all DEGs were significantly dysregulated in only one cell type, while DEGs shared across cell types showed high concordance in direction of change within major cell classes (i.e. excitatory, inhibitory, non-neuronal). Cell-type-specific schizophrenia DEGs capture transcriptomic signatures observed in bulk tissue studies, while also identifying many DEGs not detected by bulk tissue measurements. Differential gene expression within inhibitory and non-neuronal populations are captured with less efficiency by bulk tissue measurements than those within excitatory neurons, highlighting the necessity of single-cell studies.

Dysregulated genes impact biological processes within 14 broad themes relevant to brain function. Neurodevelopment-relevant processes were the most strongly enriched and primarily associated with downregulated gene sets in excitatory neurons across all cortical layers, while inhibitory neuronal populations were associated with these themes exclusively through upregulated gene sets. Synaptic signaling pathways were enriched across the greatest number of cell types and, with the exception of superficial layer excitatory neurons, associated exclusively with downregulated gene sets. The remaining ten themes were more selectively dysregulated across cell types and include many themes related to signal transduction, including “dephosphorylation,” “intracellular signal transduction,” and “cation transmembrane transport.”

These high-resolution cell-type-specific dysregulated gene sets also enabled insights into the upstream transcription factors (TFs) most likely to influence the observed expression changes. We identified a module of 24 TFs with coherent cell-type specific expression profiles strongly implicated in the regulation of schizophrenia DEGs. Multiple lines of evidence suggest a central role for the identified transcriptional regulators in schizophrenia: these TFs regulate schizophrenia dysregulated genes primarily in neurons, are genetically associated with both schizophrenia and neurodevelopmental disorders, and are key neurodevelopmental regulatory factors (95). We experimentally validated targeting of schizophrenia DEGs in neurons of the adult prefrontal cortex by a subset of these factors (TCF4, MEF2C, and SATB2), supporting our computational analysis and the relevance of the identified TFs to schizophrenia molecular pathology. This recurrent theme of neurodevelopment within TFs and biological processes emerging from analyses of schizophrenia DEGs in adult brain tissue is relevant to the neurodevelopmental hypothesis of schizophrenia (96, 97), and reflects that these regulators, classified as neurodevelopmental, also perform ongoing functions in the adult brain (98). We propose that the pleiotropic roles of these TFs, as regulators of both neurodevelopment and of neuronal schizophrenia DEGs in the adult brain, may represent a link between early neurodevelopmental disruptions and adult brain function. How genetic or early environmental perturbations to these TFs impact function to increase the risk for schizophrenia, and how they contribute to the development of prodromal symptoms and then the emergence of psychosis later in life, cannot be answered at present. However, our observations identify opportunities for future mechanistic studies tracking the developmental consequences of direct genetic perturbations to the candidate schizophrenia regulators and target genes put forward here.

We observed a significant association between schizophrenia DEGs and both common genetic variants and rare protein coding variants within excitatory populations in the superficial and deep cortical layers. Superficial layer excitatory populations showed considerable overlap in the implicated cell types, in keeping with the high similarity between the gene sets and pathways associated with these distinct categories of genetic risk (71). The strongest associations between schizophrenia DEGs and both categories of genetic risk variants occurred in excitatory neurons of deep cortical layers, and unexpectedly implicated non-overlapping populations, with Ex-L56 DEGs showing the most significant association with GWAS loci and no significant association with exome sequencing loci, and Ex-L6b_SEMA3E reversing this pattern. Outside of excitatory populations, we observed GWAS loci to be significantly linked to schizophrenia DEGs within two inhibitory populations, and exome sequencing loci to be associated with DEGs within astrocytes. If these discrepancies between the cell types most impacted by schizophrenia risk variants at opposite ends of the population frequency spectrum are replicated by future larger studies, this may suggest potential roles for these cell types in mediating distinct categories of causal genetic effects. Additionally, the observed overlap between DEGs and GWAS loci linked to multiple psychiatric disorders is consistent with the shared heritability between these illnesses (73), and such overlap was absent from Alzheimer’s GWAS loci, supporting the specificity of our analyses.

Schizophrenia is a highly heterogeneous disorder, and we assessed transcriptomic heterogeneity across individual subjects within our cohort with a measure of how consistent each individual’s transcriptional profile was with the profile characteristic of the schizophrenia group as a whole. The ranking of subjects by this measure identified two subpopulations of schizophrenia subjects, one group being transcriptionally similar to the schizophrenia group, and the other being more consistent with control subjects, an observation consistent with prior analyses of large bulk tissue RNA-seq datasets (90). While these subject groupings were not associated with subject PRS, they were well captured by the expression of two related transcriptional signatures identified across all cells in the dataset blind to subject diagnosis. These transcriptional signatures, one expressed by excitatory neuronal populations (Ex_SZCS) and the other by inhibitory populations (In_SZCS), were characterized by high expression of schizophrenia DEGs observed within multiple neuronal cell-types and by genes encoding proteins localized to the synapse.

There are multiple potential interpretations of the observed subject grouping association with these transcriptional signatures, including medication effects, compensatory changes, or genetic associations not captured by common genetic variants. We did not observe an association between medication exposures within the cohort and either subject grouping or expression of Ex_SZCS or In_SZCS, and the observation of these transcriptional signatures within multiple control subjects also argues against them being an outcome of medication exposure or compensatory change. As subject grouping and Ex_SZCS and In_SZCS appear orthogonal to PRS in this cohort, one possibility is that these transcriptional signatures identify genes influenced by non-genetic or environmental risk factors for schizophrenia. In support of such an interpretation, the gene most strongly characterizing both signatures was *DHFR*, and the fifth most associated gene with the merged signature was *CHD5*. *DHFR* encodes a key enzyme in one-carbon metabolism which impacts multiple cellular processes implicated in schizophrenia, including neurodevelopmental events, glutamatergic neurotransmission, and epigenetic mechanisms such as methylation of DNA and histone proteins (99).

Dihydrofolate reductase processes dietary folate upstream of these diverse pathways, and dietary folate supplementation during development has been associated with increased cortical thickness and decreased risk of future psychosis (100), and folate has been shown to reduce symptom severity when administered to adults with chronic schizophrenia (101). Also linking these transcriptional signatures to epigenetic processes, *CHD5* encodes a neuron specific ATP-dependent chromatin modifying enzyme within the nucleosome remodeling and deacetylation (NURD) complex that regulates gene expression through interaction with trimethylated lysine residues of histone H3 (102). During development *CHD5* acts to induce neuronal gene expression programs and is implicated in neuronal fate specification and migration (103), and it continues to be expressed throughout adulthood where its inactivation is linked to neuroblastoma (104).

This work utilized a rigorous study design and analysis plan to address potential confounds that are common to investigations of postmortem human brain tissue. Through balanced inclusion of schizophrenia and control subjects in each multiplexed sample preparation and sequencing library, we observed a remarkably low batch effect within each cohort, and very low doublet contamination, two common problems with microfluidics-based snRNA-seq, and both of these potential issues were further addressed using established computational tools. We balanced diagnostic groups for demographic variables, including age, sex, and postmortem interval, and also controlled for these variables by including them as covariates in our analyses. Our DEG analysis accounted for tissue quality by incorporating average UMI capture for each sample as a direct measure of sample performance in our experiment, influenced by known and unknown aspects of tissue integrity. Nearly all chronic schizophrenia patients receive longstanding pharmacologic treatment while individuals without psychiatric illness do not, and thus psychiatric diagnostic groups are not easily balanced for psychiatric medication exposures. We addressed this confound by performing a post hoc analysis assessing overlap of schizophrenia DEGs identified here with genes dysregulated by chronic antipsychotic exposure in non-human primates, and found no significant association.

While providing numerous biological insights, the scope of this work is limited by multiple factors. As is necessary for investigating frozen postmortem brain tissue, we measured transcriptomes of isolated single-nuclei, and the differences between nuclear and whole-tissue RNA content must be considered in the interpretation of our findings. Additionally, there are many mechanisms critical to brain function not discernible by this methodology, such as mRNA splicing or trafficking of resident dendritic mRNAs. Another technical limitation of this work is the inability to spatially resolve differential expression events, and we have addressed this problem to the extent possible through computational integration with spatial transcriptomics data. These technology-specific limitations provide an opportunity for future research when more advanced technologies become routinely available. Additionally, while our case-control design is a strength of the study, this focus on human tissue does not allow experimental manipulation to investigate the causality of our observations, and experiments in model systems are needed. Our finding of significant schizophrenia-associated perturbations within a specific subpopulation of rosehip neurons suggests that animal models may not be informative for examining some aspects of schizophrenia pathology, and other model systems, such as patient-derived induced pluripotent stem cells, may help reveal the mechanisms at play. While the direct observation of the cell type and direction of differential gene expression in disease-relevant human tissues adds valuable context to theories of disease mechanism, these data cannot determine if such changes are a cause or consequence of schizophrenia (105). Individuals with schizophrenia have very diverse experiences of treatment and clinical outcomes, and the presence of subgroups of schizophrenia subjects as suggested by our assessment of subject-specific transcriptional pathology and cellular states is promising as an analytical approach to more broadly identify such subgroups in future studies. While our combined cohort is large by present standards for single-cell transcriptional studies, replication and meaningful interpretation of these subgroups and their relation to specific environmental, genetic, and treatment factors will require assessment of larger numbers of subjects as well as additional brain regions.

Cell types were not captured with the same efficiency in both cohorts, and to characterize reproducible expression changes we restricted our primary analysis to consistently captured cell types. While this choice minimizes false-positive results, it inevitably leads to the removal of potentially relevant information. We identified several additional populations in the deeply sampled McL cohort not captured by the broadly sampled MSSM cohort which we did not analyze in depth. These include multiple excitatory neuronal subpopulations within cortical layers V and VI, including extratelencephalic projecting von Econemo neurons reported to be selectively vulnerable in schizophrenia (106, 107). Within our McL cohort, von Econemo neurons exhibited dysregulation of genes specifically enriching axonogenesis-related pathways. Similarly, our separate cohorts may be enriched for distinct biological features of schizophrenia, and focusing on the fraction of the disease-related signal that is common to both cohorts reduces false-positive associations while discarding some amount of cohort-specific true positive signal. For instance, the MSSM cohort identified oligodendrocytes as a cell-type hosting prominent schizophrenia pathology similar to previous studies (108), but this was not corroborated by the McL cohort. Cohort differences may be attributable to a variety of factors, including disease severity and chronicity. The complete datasets are available for those who wish to explore findings that did not reach our conservative thresholding for significance and reproducibility.

The data presented here complement those from others in the field (109, 110) and offer a cell-type-specific reframing of schizophrenia transcriptional pathology by revealing specific cell populations impacted by schizophrenia genes, variants, and regulators. Identification of pleiotropic transcriptional regulators linking developmental and adult schizophrenia-associated pathologies, contextualizing the outcome of known genetic risk factors within the human prefrontal cortex, and discovery of transcriptional signatures associated with heterogeneity of schizophrenia molecular pathologies provide avenues for future research to unravel the genetic and environmental underpinnings of this complex and heterogeneous disease.

## Supporting information

TableS1_SampleMetadata

TableS2_SequencingMetrics

TableS3_Celltype_Gene_Specificity

TableS4_DE_genes_merged_tables

TableS5_CrossCohortDEGoverlaps

TableS6_ReproducibleGenes

TableS7a_Down_DE_gProfiler

TableS7b_Up_DE_gProfiler

TableS8_TF_modules

TableS9_DEGvsPEG_GWAS_linkage

TableS10_CellState_Gene_Associations

TableS11_QC_Cell_Retention

## Data Availability

The source data and processed datasets described in this manuscript are available via the PsychENCODE Knowledge Portal (https://psychencode.synapse.org/). The PsychENCODE Knowledge Portal is a platform for accessing data, analyses, and tools generated through grants funded by the National Institute of Mental Health (NIMH) PsychENCODE program. Data is available for general research use according to the following requirements for data access and data attribution: (https://psychencode.synapse.org/DataAccess). For access to content described in this manuscript see: https://www.synapse.org/#!Synapse:syn25922167

## Supplementary Materials and Methods

### Assembly of the tissue cohorts

Postmortem human prefrontal cortex tissue from schizophrenia subjects and healthy control individuals matched for age and postmortem interval and including both sexes was obtained from two National Institute of Mental Health Neurobiobank associated tissue repositories, the Harvard Brain Tissue Resource Center at McLean Hospital, and The Mount Sinai/JJ Peters VA Medical Center NIH Brain and Tissue Repository. Institutional review board approval was provided by the Mass General Brigham Institutional Review Board or the Icahn School of Medicine at Mount Sinai Institutional Review Board to the relevant tissue repository for the collection, storage, and distribution of brain tissue and de-identified clinical information for each case. Diagnosis of schizophrenia or control was verified by two psychiatrists based on a review of medical records and a questionnaire completed by the donor’s family or structured interviews of informants and caregivers. Some donors within the MSSM cohort were participants in antemortem research studies. Research diagnoses based on DSM IV/V criteria were considered in reaching a final postmortem diagnosis. All cases were examined histologically, and cases meeting the criteria for known neuropathologies were excluded. Cases were obtained through family referral, and no cases were referred by a medical examiner’s office. Demographic variables for the assembled cohort are listed in Supplementary Table 1. Upon arrival at the tissue repository, fresh brains were dissected, and Brodmann Area-specific prefrontal cortex tissue (BA10 for McL, BA9, 10, or 46 for MSSM) was identified and quickly frozen with liquid nitrogen vapor or liquid nitrogen cooled isopentane prior to storage at -80°C.

### Sample processing and single-nucleus RNA sequencing (McL)

A Nissl stained cryosection from each tissue block was examined microscopically to verify that sampled tissue included all six cortical layers and underlying white matter. On dry ice 50 mg of tissue was cut from the original block and stored at -80°C until further use. Tissue samples were then processed in batches of nine (three schizophrenia, three control, and three samples not analyzed in the current study) using a protocol adapted from a previous study (29). Tissue was thawed in 0.5 ml ice-cold homogenization buffer (320 mM sucrose, 5 mM CaCl_2_, 3 mM Mg (CH_3_COO)_2_, 10 mM Tris-HCl pH 7.8, 0.1 mM EDTA pH 8.0, 0.1% IGEPAL CA-630, 1 mM β-mercaptoethanol, and 0.4 U μl−1 recombinant RNase inhibitor (Clontech)) prior to homogenization with 12 strokes in a 2 ml Wheaton Dounce tissue grinder. Tissue homogenates were passed through a 40 µm cell strainer, mixed with an equal volume of working solution (50% OptiPrep density gradient medium (Sigma-Aldrich), 5 mM CaCl_2_, 3 mM Mg(CH_3_COO)_2_, 10 mM Tris-HCl pH 7.8, 0.1 mM EDTA pH 8.0, and 1 mM β-mercaptoethanol), layered on top of an Optiprep density gradient (750 µl 30% Optiprep solution (30% OptiPrep density gradient medium,134 mM sucrose, 5 mM CaCl_2_, 3 mM Mg(CH_3_COO)_2_, 10 mM Tris-HCl pH 7.8, 0.1 mM EDTA pH 8.0, 1 mM β-mercaptoethanol, 0.04% IGEPAL CA-630, and 0.17 U μl−1 recombinant RNase inhibitor) on top of 300 µl 40% optiprep solution (35% OptiPrep density gradient medium, 96 mM sucrose, 5 mM CaCl_2_, 3 mM Mg(CH_3_COO)_2_, 10 mM Tris-HCl pH 7.8, 0.1 mM EDTA pH 8.0, 1 mM β-mercaptoethanol, 0.03% IGEPAL CA-630, and 0.12 U μl−1 recombinant RNase inhibitor)) and centrifuged at 10,000 g for 10 minutes at 4°C. Nuclei were recovered from the gradient, and resuspended in an equal volume of 1% BSA in phosphate buffered saline (PBS) prior to labeling with sample-specific cholesterol conjugated oligonucleotide hashtags (Integrated DNA Technologies). Labeled nuclei were washed with 1% BSA in PBS and pelleted by centrifugation at 500 g at 4°C for 5 minutes for three washes. After the final wash, nuclei were counted on a hemocytometer, and equal numbers from each sample were combined. The pooled nuclei were then applied to all eight channels of one 10x Genomics Chip B, targeting the recovery of 20,000 nuclei per channel. 10x Genomics and hashtag libraries were prepared as per standard 10x Genomics Chromium Single Cell 3’ Reagent Kits v3 and MULTI-Seq (111) protocols.

Libraries were sequenced in batches of two on an Illumina NextSeq500 instrument (two snRNA-Seq libraries and two hashtag libraries per flowcell - average 3.6 x 10^7^ reads per hashtag library), and an additional round of sequencing was performed for all snRNA-Seq libraries on an Illumina NovaSeq instrument (eight or 16 libraries per NovaSeq S2 flowcell) to achieve an average sequencing depth of 35,142 reads per cell. Gene count matrices were generated by aligning reads (including intronic reads) to the hg38 genome using 10x Genomics Cell Ranger software v3.0.1 (Supplementary Table 2).

### Sample processing and single-nucleus RNA sequencing (MSSM)

Six samples were processed in parallel in each batch and included a mixture of schizophrenia and control samples. All buffers were supplemented with RNAse inhibitors (Takara). 25mg of frozen postmortem human brain tissue was homogenized in cold lysis buffer (0.32M Sucrose, 5 mM CaCl_2_, 3 mM Mg(CH_3_COO)_2_, 0.1 mM, EDTA, 10mM Tris-HCl, pH 8.0, 1 mM DTT, 0.1% Triton X-100) and filtered through a 40 µm cell strainer. The flow-through was underlaid with sucrose solution (1.8 M Sucrose, 3 mM Mg(CH_3_COO)_2_, 1 mM DTT, 10 mM Tris-HCl, pH 8.0) and centrifuged at 107,000 g for 1 hour at 4°C. Pellets were re-suspended in PBS supplemented with 0.5% bovine serum albumin (BSA). 2M nuclei from each sample were pelleted at 500 g for 5 minutes at 4°C. Following centrifugation, nuclei were re-suspended in 100 µl staining buffer (2% BSA, 0.02% Tween-20 in PBS) and each sample incubated with 1 µg of a unique TotalSeq-A nuclear hashing antibody (Biolegend) for 30 min at 4°C. Prior to fluorescence-activated nuclear sorting (FANS), volumes were brought up to 250 µl with PBS and DAPI (Thermoscientific) was added to a final concentration of 1 µg/ml. DAPI positive nuclei were sorted into tubes pre-coated with 5% BSA using a FACSAria flow cytometer (BD Biosciences).

Following FANS, nuclei were subjected to 2 washes in 200 µl staining buffer, after which they were re-suspended in 15 µl PBS and quantified (Countess II, Life Technologies). Concentrations were normalized and equal amounts of differentially hash-tagged nuclei were pooled. A total of 50,400 (8,400 each) pooled nuclei were processed on a single 10x Genomics Chip B lane using single cell 3’ v3 reagents. At the cDNA amplification step (step 2.2), 1 µl 2 µM HTO cDNA PCR “additive” primer was added(112). After cDNA amplification, supernatant from the 0.6x SPRI selection step was retained for HTO library generation. cDNA libraries were prepared according to the 10x Genomics protocol. HTO libraries were prepared as previously described (112). cDNA and HTO libraries were sequenced at NYGC using the Novaseq platform (Illumina).

### Single-cell RNA data analysis

#### Quality control and cell inclusion criteria

Only cells passing a stringent, multi-step quality control protocol were considered for downstream analyses (Supplementary Table 2). Presumed doublet cells were removed using several, complementary approaches. First, the deMULTIplex R package (113) was used to process hashtag FASTQ files, extracting 10x barcode, sample hashtag, and UMI information for each read. Duplicated UMI and mismatched hashtag reads were excluded and retained reads were converted to a 10x barcode by sample, hashtag count matrix. This count matrix was processed with the Seurat R package (114) using the HTODemux function to cluster cells based on sample hashtag counts and determine a count threshold for each hashtag based on a negative binomial distribution applied to the cluster with the lowest expression for that hashtag. This threshold identified each cell as positive or negative for each sample hashtag, and cells identified as positive for more than one hashtag were assigned as inter-sample doublets and removed from the study (Supplementary Table 11). Second, we applied the doublet detection tool *Scrublet* (115) to predict and remove additional potential doublet cells. Third, genes expressed in less than 0.1% of cells, and cells with fewer than 1,000 identified genes, more than 10% of unique molecular identifiers (UMIs) in mitochondrial genes, or more than 50,000 captured UMIs were removed. The previous three steps were applied to each batch independently. Finally, an additional QC step was applied during cell annotation and clustering (see below). Outlier cells and cell clusters defined by having extreme values of quality control measures (total UMI count, percentage of reads mapping to mitochondrial genes) and extremely low similarity/connectivity with other cells of the same type were empirically identified, manual examined in exploratory 2D projection plots, and removed from downstream analyses.

#### Cell annotation and clustering

All normalization, data integration, dimensionality reduction, clustering, annotation, visualization, and matrix decomposition-based analyses were performed using our computational environment for single-cell analysis ACTIONet (39). Briefly, ACTIONet initially performed single value decomposition (SVD) for feature (gene) dimensionality reduction and selection, producing a low-rank approximation of the normalized count matrix, which is subsequently analyzed using archetypal analysis to define a low dimensional representation for each individual cell transcriptome. This final archetypal decomposition is used to build a cell network (manifold) capturing relationships in transcriptomic state similarity at single-cell resolution. Cells from all batches and cohorts were analyzed jointly using ACTIONet’s integrated batch correction functionality based on Harmony (116) to remove potential technical effects associated with different batches and/or cohorts. Subsequently, a cell-cell similarity network was constructed based on the batch-corrected and lower dimensional data. Next, a curated set of reproducible cell-type-specific markers (39) were used to annotate the major cell types of the human cortex based on cellular-level expression patterns, as implemented in the *annotate.cells.using.markers* function of ACTIONet’s R package (https://github.com/shmohammadi86/ACTIONet/tree/R-release). To further dissect cellular diversity and identify low-abundance cell types, neuronal subtypes, and remaining low-quality cells, cells associated with each major cell type were reanalyzed separately using the same ACTIONet-based procedure followed by graph clustering. The resulting clusters were subsequently annotated using the known cortical layer (40) and cell type markers (29, 30, 39) from the literature. A final set of cell labels was defined by integrating higher-resolution annotations inferred during cell type clustering analyses with previous cell type level annotations. All annotations and interpretations were corroborated empirically by visualizing the expression patterns of individual marker genes across cells in 2D projection plots.

### In situ hybridization

Three frozen tissue blocks of the prefrontal cortex, Brodmann area 10, from unaffected control donors were obtained from the HBTRC. The blocks were embedded into optimal temperature cutting medium (OCT) and frozen at -20°C. 10 μm sections were cut with a NX 70 cryostat (Thermofisher) at -20 °C, collected onto Superfrost Plus Gold slides and stored -80°C until Duplex chromogenic in situ hybridization (ISH) was performed.

Advanced Cell Diagnostics (ACD) designed the in situ hybridization probes (GAD1 Probe ID 404031, Channel 1, Enzyme Horseradish Peroxidase (HRP); SV2C Probe ID 448361, Channel 1 Enzyme HRP and Channel 2 Enzyme Alkaline Phosphatase (AP); TRPC3 Probe 427641, Channel 1, Enzyme HRP; CHST9 Probe ID 1096741, Channel 2, Enzyme AP) as well as positive and negative control probes. The RNAscope 2.5 HD Duplex detection kit (Chromogenic, # 322500, ACD) was used for the assay which was carried out following the manufacturer’s instructions with some modifications. For detection of the signal, Fast Red, and Green substrates (ACD) were used. Briefly, frozen sections were fixed for 1 h at 4 °C using freshly prepared ice-cold 10% neutral buffered formalin. Afterward, sections were rinsed with phosphate buffered saline (PBS) and dehydrated in 50 %, 70 %, and two changes of 100 % ethanol (5 min each) at room temperature (RT). Sections were air-dried and a hydrophobic barrier drawn around each section with an Immedge pen (Vector Laboratories). When completely dry, sections were treated with hydrogen peroxide for 10 min at RT, washed twice with PBS, incubated with protease IV for 15 min at RT, and washed again twice with PBS.

After diluting the probes for RNA detection (1:50), sections were hybridized with the probes (40°C, 2 h; HybEZ Hybridization System (ACD)), washed twice and stored overnight at RT in 5x SSC buffer. On the next day, slides were washed twice with wash buffer, followed by ten amplification (AMP) steps. Each AMP step was followed by two washes of 2 min each with wash buffer. After the AMP 6th step, sections were incubated with the Fast Red substrate to detect the red signal (e.g. AP-C2 probes; RT, 10 min). At the end of AMP 10, sections were incubated with Green substrate to detect the green signal (e.g. HRP-C1 probes; RT 10 min). Gill’s Hematoxylin I (50%) was used (30s) to visualize cell nuclei. Sections were mounted using VectorMount medium (Vector Labs # H 5000) and stored at RT. All microscopy was performed at the Microscopy CoRE at the Harvard Center for Biological Imaging. Imaging was performed on an Axio Scan.Z1 automated slide scanner using a 20X (0.8NA) objective. Brightfield images were captured on a Axiocam 705 CMOS camera and images were processed with ZEN 3.5 blue edition software.

### Differential gene expression analysis

Differential expression (DE) analysis was performed using standard protocols implemented in the Multi-sample multi-group scRNA-seq data analysis tools (*muscat*) R package (117). All analyses were conducted using pseudobulk gene expression profiles with log-transformed expression counts (see data availability) and the method limma-trend, which fits a linear model to aggregate expression profiles to estimate and quantify gene expression changes. Outlier genes were filtered out using *muscat’s* default criteria. Only the 25 cell types captured (representing at least 5 cells) within a total number of subjects larger than the number of covariates were considered for the analysis. To minimize biases due to differences in cell composition, DE analysis was performed using only those samples showing consistent cell-type proportions as identified in the cell-type annotation step (135 subjects total; McL 21 schizophrenia and 24 control, SZ3, SZ15, and SZ24 excluded; MSSM 39 schizophrenia and 51 control, SZ29 and SZ33 excluded; exclusions based on excitatory neuronal populations accounting for greater than 80% or less than 10% of total cells; (**Extended Data Figure 1f**). DE analyses were performed separately for each cohort and cell type, after removing the effect of batch and HTO variables using *removeBatchEffect* function in Limma (118), while incorporating the following covariates: subject age, sex, postmortem interval, and the log transform of the average number of UMIs captured per cell (Supplementary Table 1). UMI capture was included as a direct assessment of sample performance to control for known and unknown potential confounds related to sample quality and processing.

#### DE meta-analysis

Meta-analysis was performed over all genes via a fixed effects meta-analysis (119) in each of the 25 cell types robustly captured in each tissue cohort. FDR multiple testing correction was performed over all combined genes in each cell type independently. DE genes were selected based on the following criteria: FDR-corrected p-value < 0.05 and absolute logFC > 0.1.

### Functional enrichment of DE genes

Overrepresentation tests for GO biological processes for all 50 schizophrenia DEG sets (upregulated and downregulated gene sets tested independently) were performed using the R package gprofiler2 (120). To facilitate interpretation of DE results, a coherent and non-redundant set of *meta*functional *terms* was defined by grouping GO biological processes based on their semantic similarity using the R package rrvgo, which resulted in the identification of 14 interpretable categories.

### Quantitative polymerase chain reaction validation of differential gene expression

Total RNA from the frozen prefrontal cortex, Brodmann area 10 tissue was extracted with TRIzol Reagent (Thermo Fisher Scientific #15596018) and RNeasy Miniprep Kit (Qiagen 74104). Briefly, 50-75 mg of tissue was homogenized in 800 μL of TRIzol using pellet pestles (Sigma-Aldrich ref: Z359947) coupled to pellet pestles cordless motor (Sigma-Aldrich ref: Z359971) in 1.5 ml tubes. Once homogenized, samples were centrifuged for 5 min at 16,000 g at 4 °C to remove any remaining tissue pieces and the supernatant was transferred to clean 1.5 mL tubes. 200 μL of chloroform was added, tubes were vortexed for 5 sec and left at RT for 10 min. To obtain the phase separation, samples were centrifuged for 15 min at 10,000 g at 4 °C. The aqueous upper face containing the RNA was transferred into a clean Eppendorf tube and RNA was extracted with RNeasy Miniprep RNA Kit following manufacturer’s instructions and including DNase I on column treatment. RNA was quantified by measuring absorbance at 260 nm using a Nanodrop ND-100 (Thermo Fisher Scientific).

cDNA was generated from 2 µg RNA per sample using iScript Advanced cDNA synthesis kit (# 1725037 Bio-Rad). PCR reactions were run on a CFX96 Touch Real Time PCR (Bio-Rad) with SsoAdvanced™ Universal SYBR® Green Supermix (# 1725270 Bio-Rad). qPCR primer pairs were obtained from Integrated DNA Technologies (B2M - HA.PT.58v.18759587; BCR - Hs.PT.56a.2126741; BSN - Hs.PT.58.3309612; NEURL1B - assay Hs.PT.58.425664.g; PSEN1 - Hs.PT.58.38987623; RASGRF1 - Hs.PT.58.875114; SHANK2 - Hs.PT.58.807139.g; SLC1A2 - Hs .PT.58.26435316). Real-time PCR reactions were carried out using 4 µl (1:10 diluted) of cDNA for each reaction in a 20µl volume following the manufacturer’s protocol. For each sample and gene, three replicates were run in a 96-well plate. The housekeeping gene β-2-microglobulin was used as an endogenous control. Fold change of expression of each target gene in each schizophrenia subject relative to average expression across all control subjects normalized to 1 was computed with the ΔΔct method and beta-2 microglobulin (B2M) used as the reference gene.

### Identification of putative transcriptional regulators

The ChIP-X Enrichment Analysis 3 (ChEA3) tool (62) was used to prioritize transcription factors (TFs) based on the overlap of their putative targets and the set of genes observed to be significantly transcriptionally dysregulated (FDR < 0.05 and log(fold change)>0.1)) in schizophrenia. ChEA3 integrates multiple libraries of putative TF-target lists, gathered from multiple sources, including TF-gene co-expression from RNA-seq studies, TF-target associations from ChIP-seq experiments, and TF-gene co-occurrence computed from crowd-submitted gene lists. The *TopRank* strategy was used to combine TF rankings from individual libraries. In this strategy, the best scaled-rank of each TF across all libraries is used to aggregate TF scores. In the present study, the log-transformed scaled-rank was used as the relevance measure for a given TF according to ChEA3 enrichment analysis.

### Computing transcription factor co-expression modules

To identify groups of transcriptional factors with similar expression profiles across cells (TF modules), weighted correlation network analysis (WGCNA) was used to construct gene co-expression modules between transcription factors across cell types (121). Pseudobulk profiles were used to construct an independent network for each cell type. For each cell type, the unsigned co-expression similarity measure with β = 6 and transformed similarity scores using the topological overlap measure (TOM) was used as a measure of proximity. The 95% percentile of each proximity score was then matched and averaged across cell types to compute a consensus similarity matrix. Finally, hierarchical clustering with the average (UPGMA) agglomeration method was applied to the consensus matrix to obtain the final TF modules. This resulted in the identification of the 16 TF modules used in the study. The resulting TF modules were then scored by the average of the individual ChEA3 scores obtained for the members of the module. Only one TF module showed a high average ChEA3 score.

### CUT&Tag mapping of transcription factor binding in the neuronal genome

Nuclei were isolated from the postmortem human prefrontal cortex as described above and incubated with 1:1000 diluted anti-NeuN antibody (EMD Millipore MAB377X) with 0.5% BSA in PBS at 4°C with end-over-end rotation for 45 minutes. After staining, nuclei were counterstained with propidium iodide and sorted on a BD FACSAria III Cell Sorter at Harvard University’s Bauer Core Facility. 100,000 neuronal nuclei were used as input for each Cleavage Under Targets and Tagmentation (CUT&Tag) assay using rabbit primary antibodies targeting MEF2C (Abcam ab211493), SATB2 (Abcam ab92446), TCF4 (ProteinTech 22337-1-AP), NFKB (Proteintech 14220-1-AP), and mouse-anti-rabbit secondary antibody (Sigma R2655) with the Vazyme pG-Tn5 CUT&Tag kit (Cellagen Technology, San Diego) according to the manufacturer’s protocol. CUT&Tag libraries were sequenced on one NextSeq500 flow cell at the MIT BioMicroCenter.

### CUT&Tag

*Computational analysis*: Sequenced reads were aligned to the HG38 genome, processed to bedgraph format, and CUT&Tag signal enrichment was assessed by peak calling using the Sparse Enrichment for CUT&Run (122) tool with “stringent” parameters and the top 1% (fdr<=0.1) of peaks were considered as genomic elements with evidence of binding. To investigate the functions of genes being targeted in our neuronal sorted samples, a master-set of bound regions was defined for each TF by considering all regions with evidence of binding across samples. The resulting regions were annotated with their nearest gene, filtering out those farther than 100k bps, and the resulting genes were subjected to pathway enrichment analysis using the ReactomePA and clusterProfiler R packages. To assess the overlap of TF binding and Sz DEGs a more stringent set of reproducibly bound events was defined by requiring binding support in at least 50% of samples, and the overrepresentation of DEGs within the genes annotated to these reproducibly bound regions was tested using the Fisher’s exact test considering only genes with detected expression in the corresponding cell type as background. All region annotations were performed using the ChIPseeker R package (123).

### Comparison of GWAS risk and differential expression

The computational tool Hi-C-coupled MAGMA (H-MAGMA (72)) was used to assess associations between schizophrenia DE genes and genetic risk genes of representative brain disorders. H-MAGMA is a recent extension of the multimarker analysis of genomic annotation (MAGMA), a method developed to prioritize GWAS genes by aggregating single nucleotide polymorphism associations to nearest genes. In H-MAGMA, the linking of SNPs to genes is extended to include long-range interactions based on evidence provided by chromatin conformation capture Hi-C experiments. All analyses presented herein are based on H-MAGMA’s geneset enrichment approach using the preprocessed dataset of SNP-gene links generated by Hi-C experiments in the adult human brain (65).

### Transcriptional pathology score (TPS)

To assess the degree of schizophrenia-associated transcriptional changes within individual subjects, as a continuous measure of transcriptional dysregulation in schizophrenia complementing the binary case-control comparison DEG analysis, we introduced a quantitative measure of transcriptional pathology based on cell-type-specific pseudobulk profiles. For each cell type, we first compute a mean gene expression pattern across all subjects. We then computed the partial Pearson’s correlation between the expression profile of each subject and the differential expression profile observed between the schizophrenia and control groups in our cross-cohort DEG meta-analysis, after correcting for the baseline (mean) expression of genes in the given cell type. This produced an individual by cell type TPS matrix, and a donor level TPS was defined as the average of TPS across all neuronal cell-types for each individual (**Fig. 4a**).

### Decomposition-based cell state discovery

Cell state analysis was performed using our computational analysis framework ACTIONet (39) available at (https://github.com/shmohammadi86/ACTIONet/). Briefly, to identify a set of dominant transcriptional patterns explaining the cellular heterogeneity in the dataset, a single value decomposition (SVD)-based preprocessing step is first performed for dimensionality reduction, producing a low-rank approximation of the normalized count matrix. This reduced data representation is subsequently decomposed multiple times using an archetypal analysis (AA)-based approach with increasing resolution (number of archetypes) to define a multiresolution and low-dimensional cell state representation for each individual cell. This approach is supported by the recent finding that archetypal analysis of the SVD-reduced profiles closely approximate the archetypes of the original dataset while reducing noise and computational burden (124). The default settings, as implemented in the ACTIONet R package function *runACTIONet,* were used for all decompositions in the present study. This procedure results in the decomposition of an input normalized count matrix into a matrix (W) of archetypes (cell states) and a matrix (H) of cell encodings. The first matrix represents cell states in terms of gene expression profiles, while the latter encodes the contribution of a given transcriptional state to the transcriptome of each cell. The encoding matrix H was used to assess the degree to which archetypes recover cell types using overrepresentation analysis of cell type labels. This procedure allows discriminating between cell type-associated archetypes and those recovering patterns shared across cell types. The majority of identified archetypes were closely aligned with annotated cell types, with the exception of four archetypes that exhibit cross-cell-type variations including one archetype (In_SZCS) that strongly differentiated subgroups within diagnoses.

### Genetic ancestry and polygenic risk score analysis

#### DNA preparation and SNP array processing (MSSM)

DNA was isolated and genotyping was performed as previously described (44). Briefly, all DNA isolation was done from approximately 10 mg dry homogenized tissue using the Qiagen DNeasy Blood and Tissue Kit according to the manufacturer’s protocol. DNA yield was quantified using Thermo Scientific’s NanoDrop. The mean yield was 12.6 ug (±4.6 ug), the mean ratio of 260/280 was 2.0 (±0.1) and the mean ratio of 260/230 was 1.9 (±2.3). Genotyping was performed on the Illumina Infinium HumanOmniExpressExome 8 v 1.1b chip (Catalog #: WG-351-2301) using the manufacturer’s protocol as previously described (125). Normalized bead intensity data obtained for each sample were called using Illumina Genome Studio with cluster position files provided by Illumina, and fluorescence intensities were converted into SNP genotypes.

#### DNA preparation and SNP array processing (McL)

For each donor DNA was isolated from 50 mg of postmortem human brain tissue using the Qiagen DNeasy Blood and Tissue Kit according to the manufacturer’s protocol. DNA yield was quantified using the Quant-IT PicoGreen dsDNA Assay Kit and genotyping was performed on the Illumina Infinium HumanOmniExpressExome 8 v 1.6 chip (Catalog #: 20024677) by the Broad Institute’s Genomics Platform according to the manufacturer’s protocol. Normalized bead intensity data obtained for each sample were called using Illumina Genome Studio with cluster position files provided by Illumina, and fluorescence intensities were converted into SNP genotypes.

#### Imputation

Imputation was performed separately for each cohort. Markers were lifted-over to GRCh38 and and aligned to the TOPMed version R2 (126) loci using HRC-1000G-check-bim-v4.3.0 (https://www.well.ox.ac.uk/~wrayner/tools/), which checks the strand, alleles, position, reference/alternate allele assignments and frequencies of the markers, removing A/T & G/C single nucleotide polymorphisms (SNPs) with minor allele frequency (MAF) > 0.4, SNPs with differing alleles, SNPs with > 0.2 allele frequency difference between the genotyped samples and the TOPMed samples, and SNPs not in the reference panel. Imputation was performed via the TOPMed Imputation Server (https://imputation.biodatacatalyst.nhlbi.nih.gov/) which is based on previously developed software (127) using Eagle (128) as the haplotype phasing algorithm. Imputed SNPs from both cohorts with imputation R^2^ > 0.3 were retained for downstream analyses.

#### Relatedness and population stratification

All individuals in this study were unrelated (kinship coefficient cut-off of ≥ 0.0884). Relatedness was estimated by robust relationship inference algorithm (129) using Plink v2 (130) after LD-based pruning of common variants. Genotypes from the 1000 Genomes Project were lifted-over to GRCh38 and merged with the imputed genotypes from McLean and MtSinai; variants were filtered (MAF ≥ 0.01, Hardy-Weinberg P value > 1 × 10^−10^), pruned, and used for PCA analysis with Plink v2 (130). A three-dimensional ellipsoid based on the first three PCs of the reference 1000G superpopulation (e.g. Europeans) with a diameter of three standard deviations was created and those who fell outside this ellipsoid were considered as non-members (e.g. non-Europeans) and excluded from the analysis.

#### Schizophrenia polygenic risk score (PRS) calculation

Polygenic scores were constructed for schizophrenia using effect sizes from the largest to-date schizophrenia GWAS (15). The summary statistics were processed using PRS-CS software (131) to generate weights (posterior SNP effect sizes). Default settings were used for calculating weights using PRS-CS (γ-γ prior=1; parameter b in γ-γ prior=0.5; MCMC iterations=1000; number of burn-in iterations=500; thinning of the Markov chain factor=5). Then, based on the derived weights individual level polygenic scores for schizophrenia were calculated using Plink v2 (130) software in a curated set of SNPs from the imputed genotypes (imputation R^2^ > 0.8, MAF ≥ 0.01; Hardy-Weinberg P value < 1 × 10^−10^). Schizophrenia PRS was scaled (mean=0, sd=1) in the individuals of European descent in the combined cohort.

## Acknowledgements

We thank the individuals and families whose donation of human brain tissue made this work possible. This work was supported by the Wilf Family Foundations and by NIH grant K08MH109759 (W.B.R.), by NIH grant K08MH122911 (G.V.), by NIH grant HHSN271201300031C (V.H.), by NIH grants R01AG050986, R01MH109677, U01MH116442, R01MH110921, R01MH125246 and R01AG067025 and the Veterans Affairs Merit grant BX002395 (P.R), and by NIH grants 1U01MH119509, R01MH109978, and R01AG062335 (M.K.).

## Author Contributions

This study was designed by W.B.R., J.F.F., and P.R., and directed and coordinated by W.B.R., P.R., and M.K. V.H. provided tissue dissections (MSSM). W.B.R., S.S., and D.R.T. (McL) and J.F.F (MSSM) performed the snRNA-seq experiments, and W.B.R. and S.S. performed the CUT&Tag experiment. S.S. performed the RNAscope and qPCR experiments. W.B.R. (McL) and S.J. and H-C.L. (MSSM) performed data processing and S.M. and J.D.-V. performed the computational analysis. G.E.H. assisted with data integration and meta-analysis. G.V. performed the genetic analyses. D.R.T. and M.H. reviewed medical records under supervision of W.B.R. W.B.R., S.M., J.D.-V., J.F.F., G.E.H., P.R., and M.K. wrote the manuscript.

Data were generated as part of the PsychENCODE Consortium. Visit https://www.synapse.org/#!Synapse:syn24240356 for a complete list of grants and PIs.

## Supplemental Figures

**Extended Data Fig. 1a.**
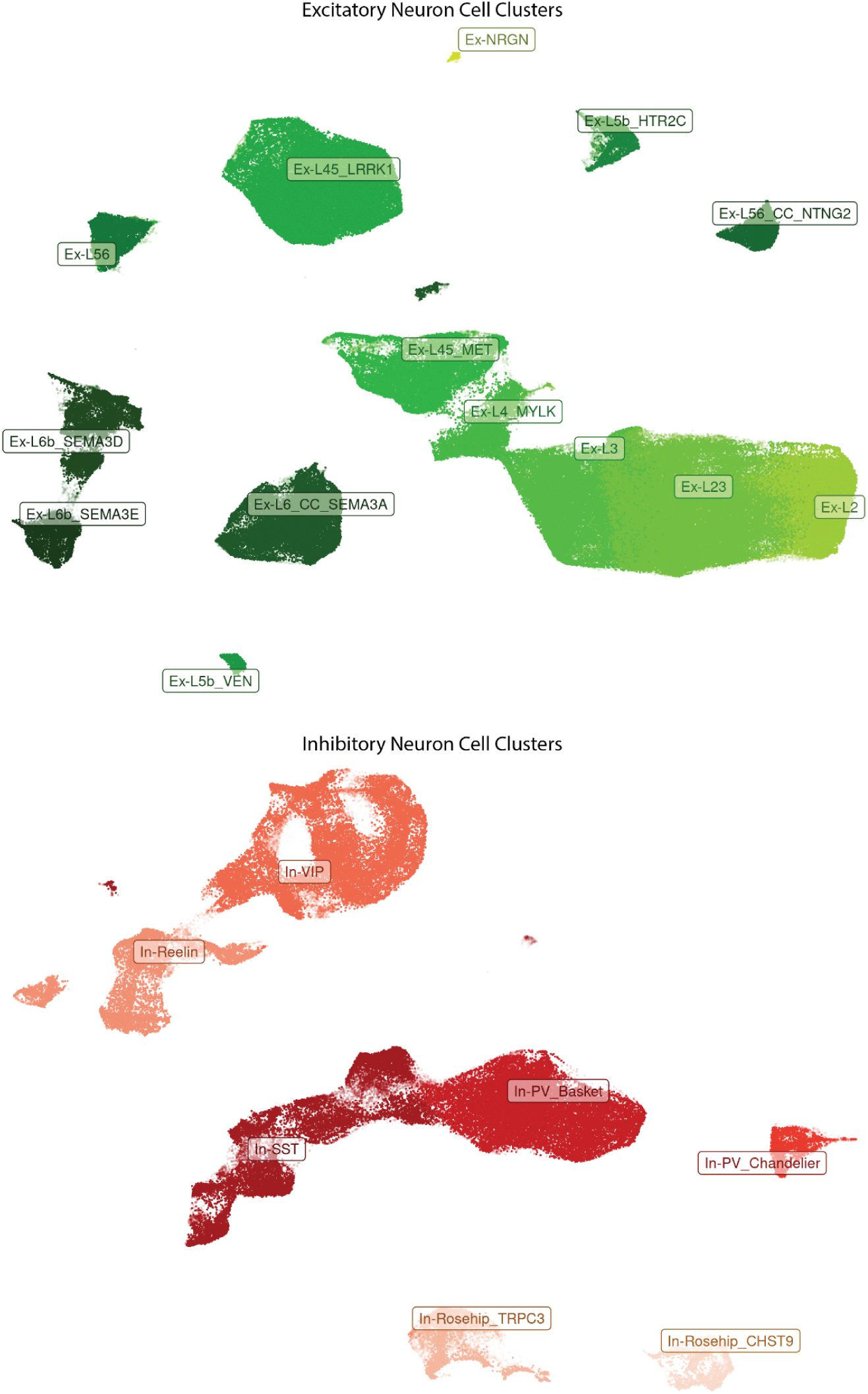
Subclustering of neuronal cells identifies 14 distinct excitatory (top) and seven inhibitory (bottom) populations.

**Extended Data Fig. 1b, c, d, e.**
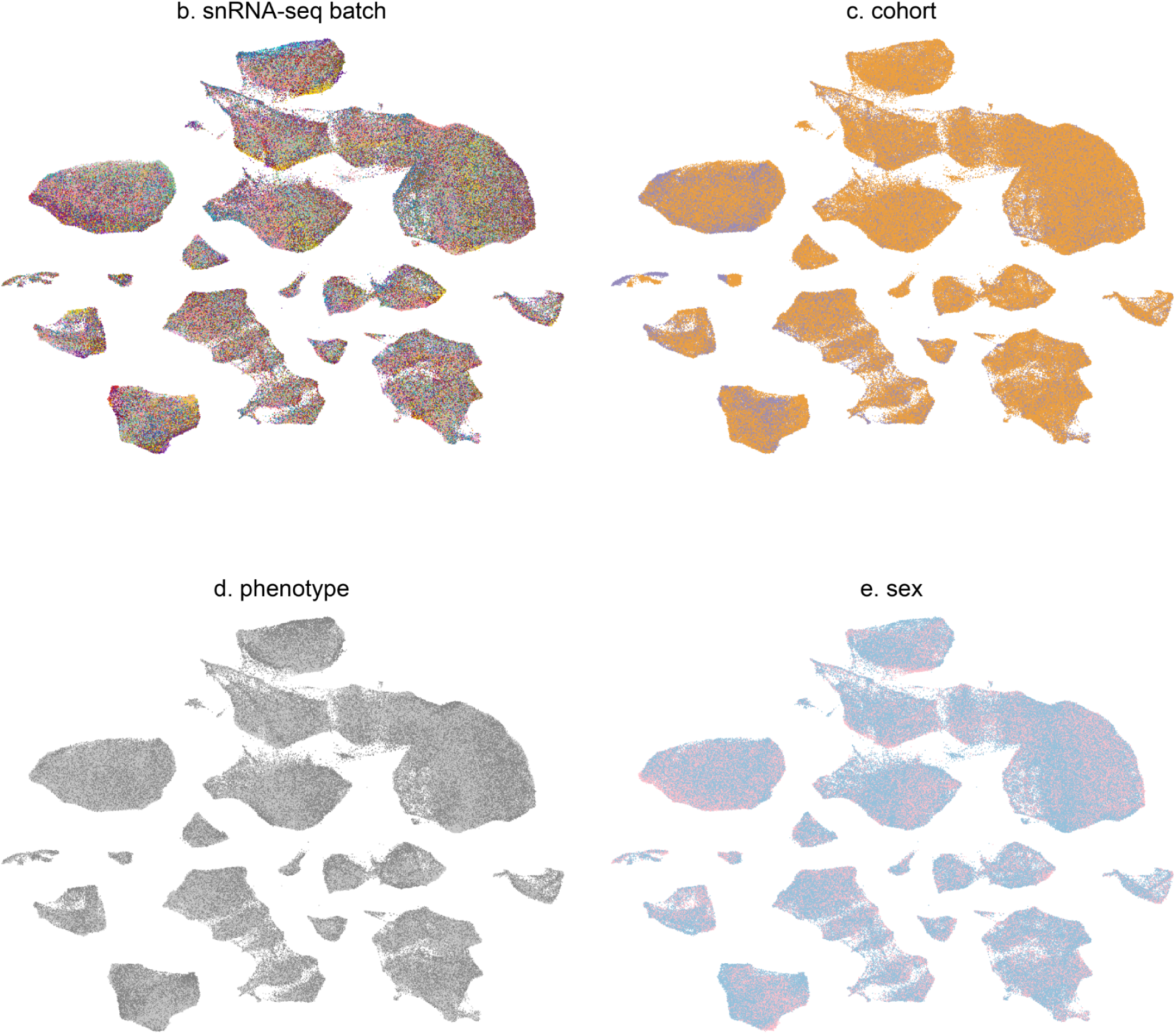
ACTIONet cell-cell similarity network. Network-based two-dimensional visualization of all cells considered in the analysis (n=468,727) depicting clustering of cells within transcriptomically identified distinct cellular populations showing cells colored by snRNA-seq batch (b), cohort (c - orange:McLean, n=361,966; blue:Mount Sinai, N=106,761), phenotype (d - dark grey: schizophrenia, n=206,014; light grey: control, n=262,713), or sex (e - red: female, n=141,194; blue:male, n=281198), demonstrating even mixing and contribution from batches, cohorts, phenotypes, and sexes within each major cell population.

**Extended Data Fig. 1f, g.**
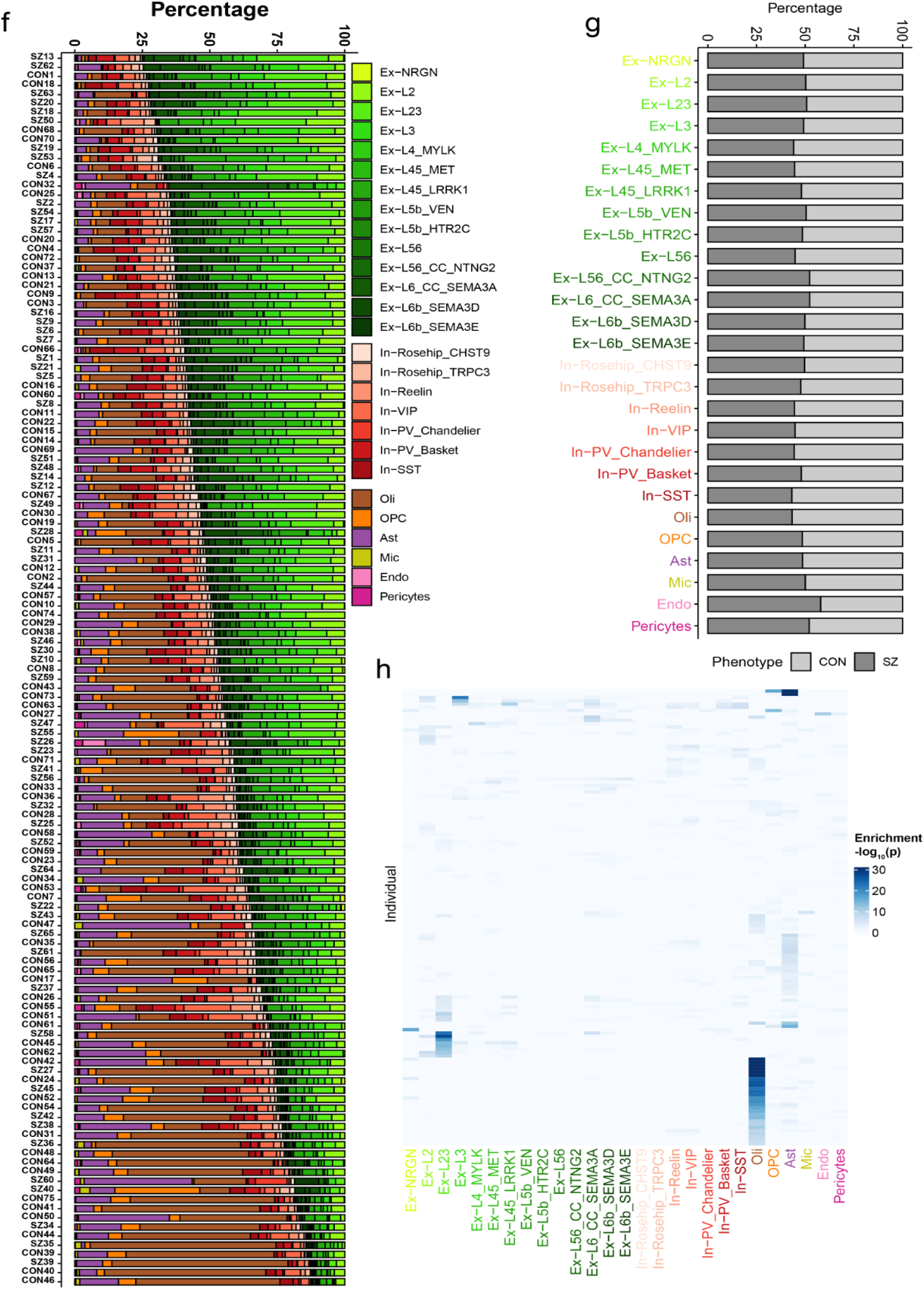
Cell-type fractions across individuals and diagnostic groups. **f.** Cell type fractions across individuals. **g.** Cell type fractions across diagnoses demonstrating no enrichment of cell types within schizophrenia or control groups. **h.** Cell type enrichment within individuals showing overrepresentation of oligodendrocytes within some samples without appreciable enrichment of other cell types.

**Extended Data Fig. 2a.**
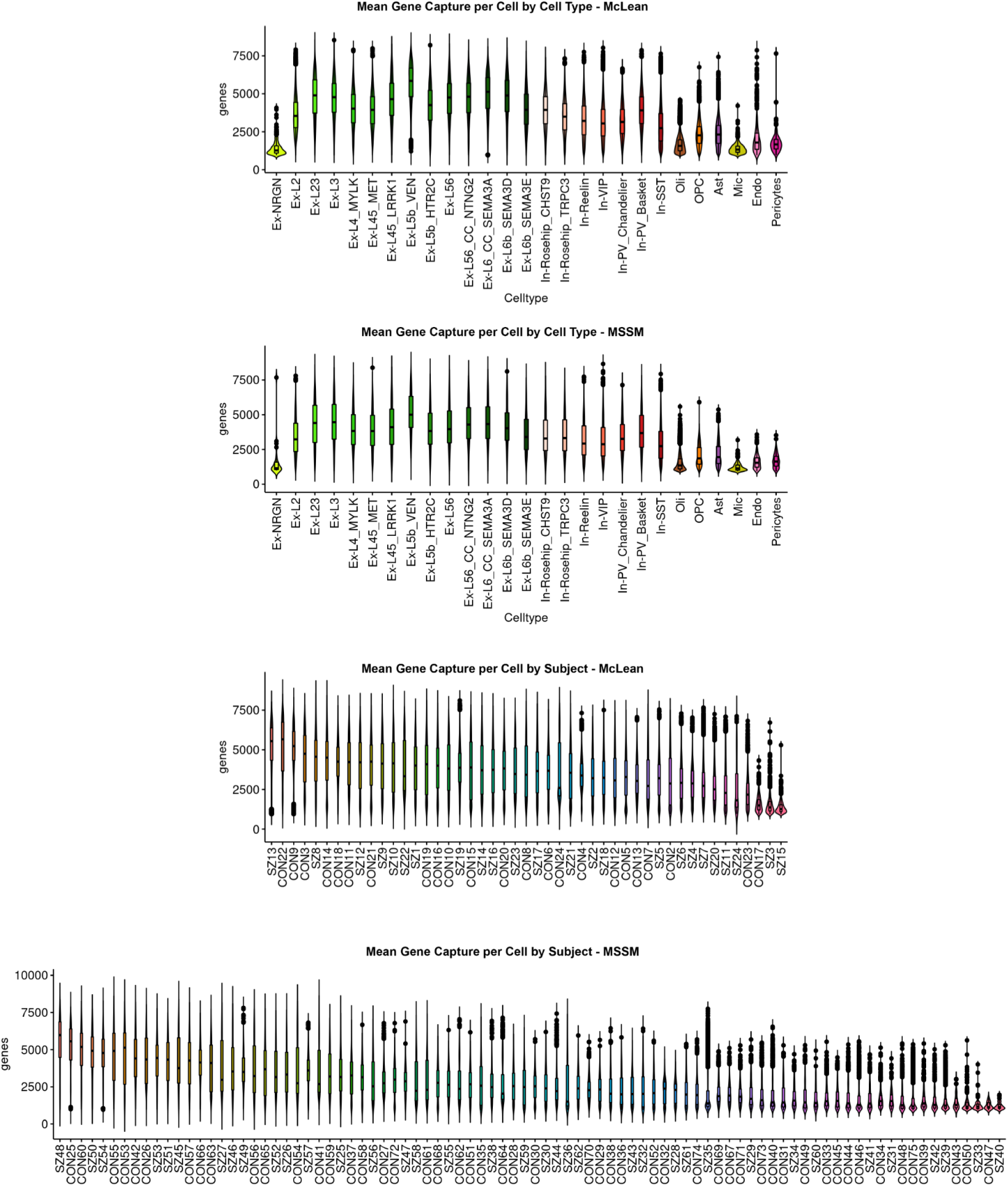
Gene capture across cell-types and individuals. Gene count distribution across cells of each type. Each point represents in log scale the number of genes detected to have a read count x > 0 in a given cell or individual for each cohort. Box plots are centered around the median, with the interquartile range (IQR) defining the box. The upper whisker extends to the largest value no further than 1.5 × IQR from the end of the box. The lower whisker extends to the smallest value at most 1.5 × IQR from the end of the box.

**Extended Data Fig. 2b.**
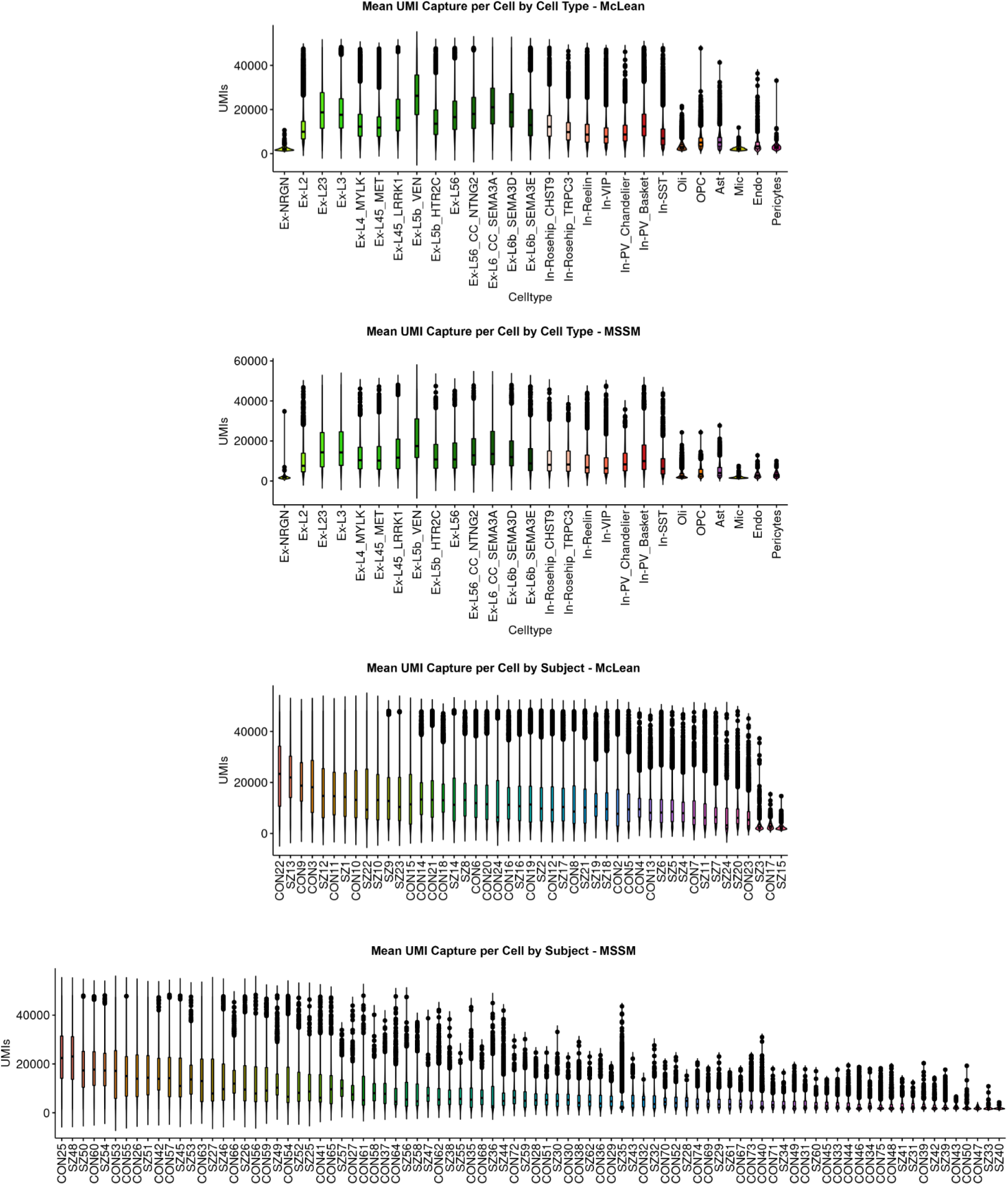
UMI capture across cell-types and individuals. Total UMI count distribution across cells of each type. Each point represents the total UMI count across all genes in a given cell or individual for each cohort. Box plots are centered around the median, with the interquartile range (IQR) defining the box. The upper whisker extends to the largest value no further than 1.5 × IQR from the end of the box. The lower whisker extends to the smallest value at most 1.5 × IQR from the end of the box.

**Extended Data Fig. 2c.**
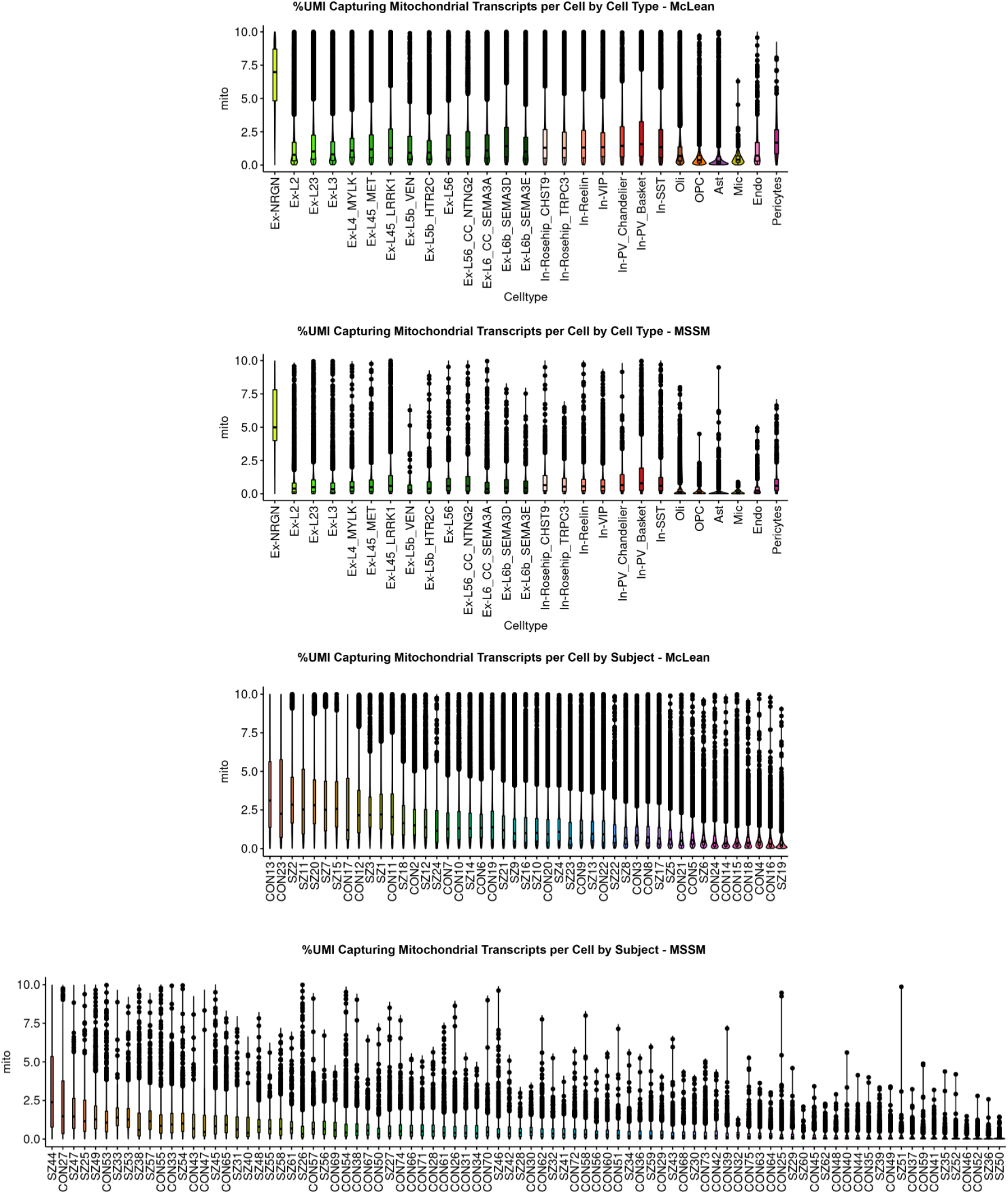
Mitochondrial transcript capture across cell-types and individuals. Distribution of UMI percentages that map to mitochondrially encoded genes across cells of each type or individual for each cohort. Box plots are centered around the median, with the interquartile range (IQR) defining the box. The upper whisker extends to the largest value no further than 1.5 × IQR from the end of the box. The lower whisker extends to the smallest value at most 1.5 × IQR from the end of the box.

**Extended Data Figure 3.**
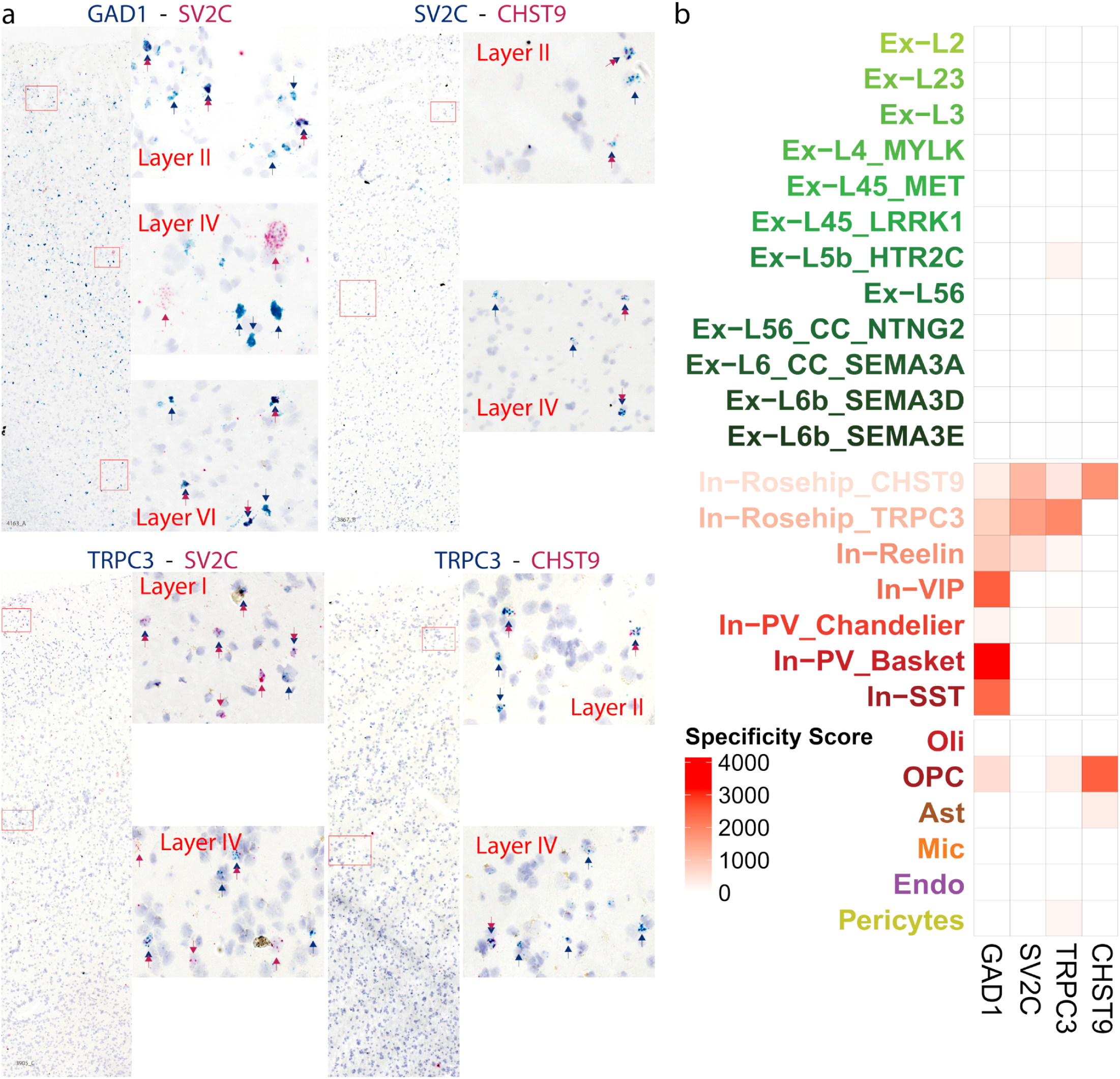
In situ hybridization validation of Rosehip interneuron heterogeneity. **a.** Duplex chromogenic RNAscope staining of pairwise combinations of four mRNAs was performed in 10 um thick sections of postmortem human Brodmann Area 10 showing good agreement with the expected colocalization of these transcripts predicted by our snRNA-seq data. As expected, SV2C, a marker of the Rosehip interneuron subtype, is expressed primarily by a subset of GAD1 expressing interneurons, with some SV2C expression unexpectedly observed within GAD1 negative cells. CHST9 was expressed within a subset of SV2C positive cells, with very few observations of CHST9+/SV2C- cells. Similarly, TRPC3 was expressed by a subset of SV2C positive cells, with more observations of TRPC3+/SV2C- cells as expected. Finally, CHST9 was expressed by a subset of TRPC3 positive cells, identifying a subset of transcriptionally distinct Rosehip interneurons. Throughout all panels, colored arrowheads point out cells labeled with only the green or red probe, and double arrowheads indicate double labeled cells. **b.** Heatmap depicting the specificity of expression across cell types of the four transcripts stained for in panel a used to predict overlapping labeling patterns. Specificity scores are computed using one-sided Wilcoxon rank-sum test. Specificity scores are computed by the -log(pvalue) from preferential expression tests implemented in ACTIONet.

**Extended Data Fig. 4.**
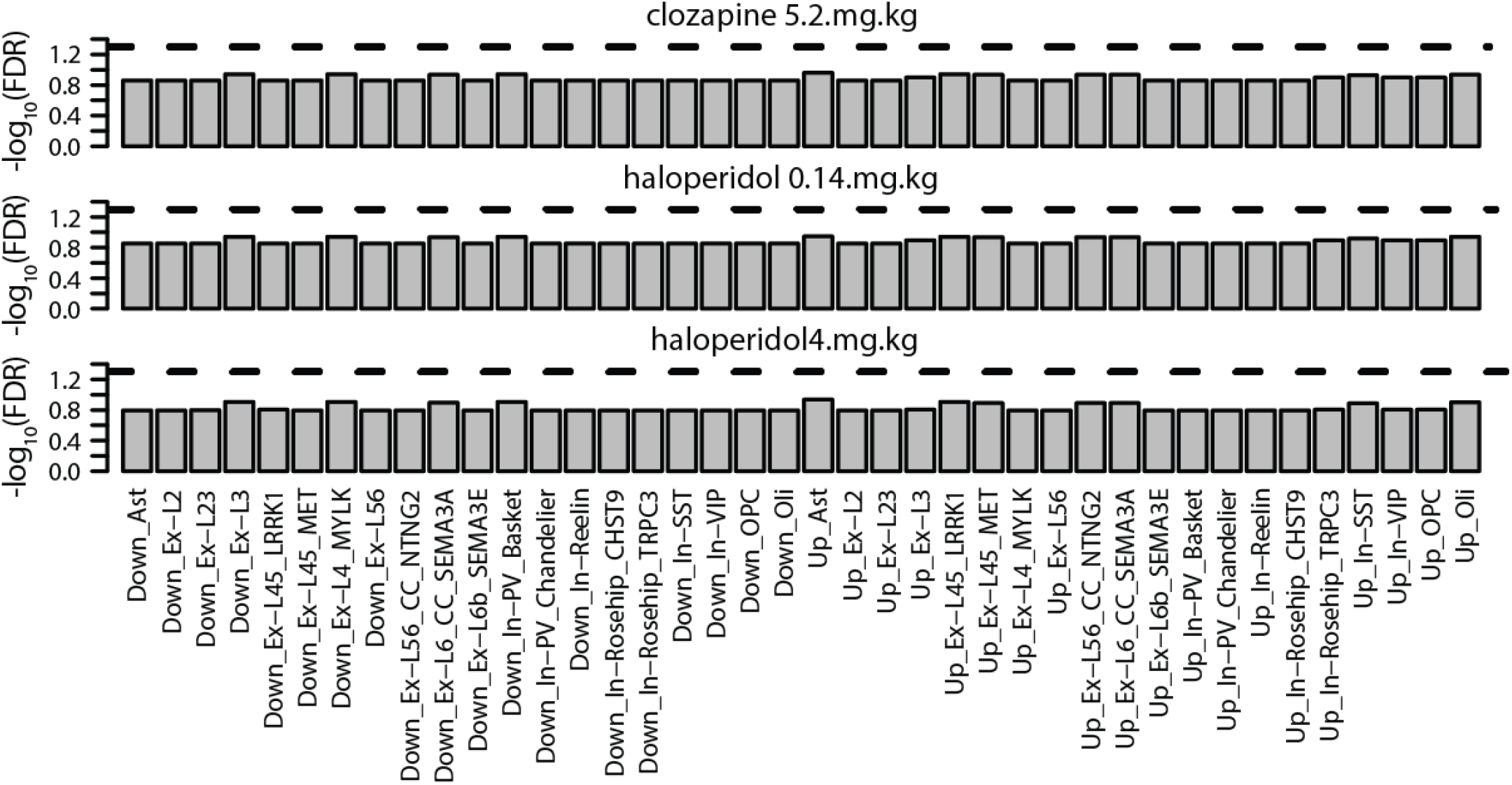
Genes affected by chronic antipsychotic exposure are not enriched for cell-type-specific down or upregulated DEGs. Post hoc analysis of cell-type-specific DEG sets find no significant overlap with genes that are dysregulated in non-human primates in response to chronic treatment with clozapine or with high or low dose haloperidol.

**Extended Data Fig. 5.**
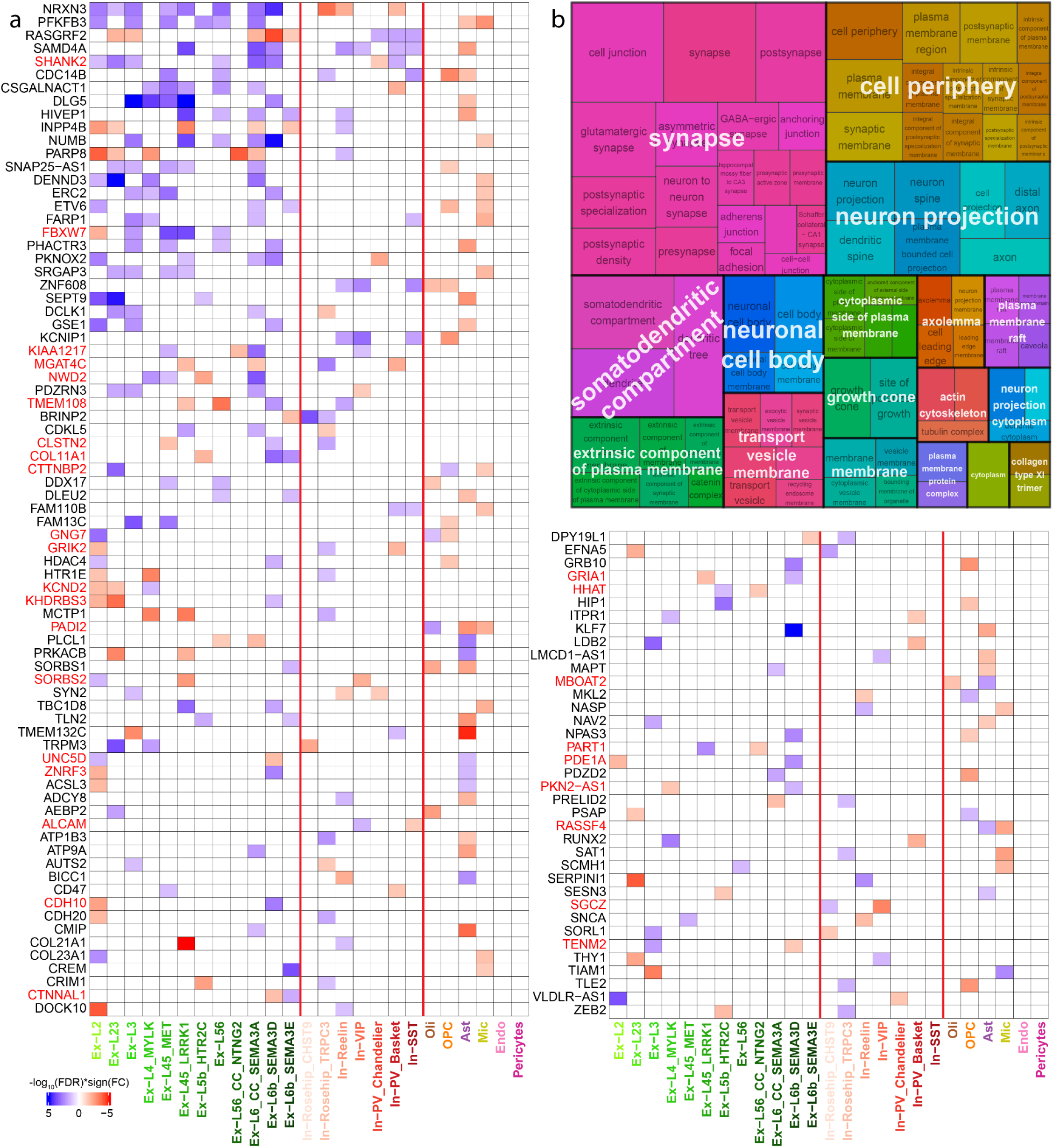
**a.** Differential expression of all 114 divergently dysregulated schizophrenia DEGs across cell types. The 29 genes that violate the trend of concordant dysregulation within cell classes are named in red. Heatmap is split only for space considerations. **b.** Treemap of semantically clustered Gene Ontology Cellular Component terms significantly enriched (FDR < 0.05) by the 114 divergently dysregulated schizophrenia DEGs demonstrating abundance of synapse related terms.

**Extended Data Fig. 6a.**
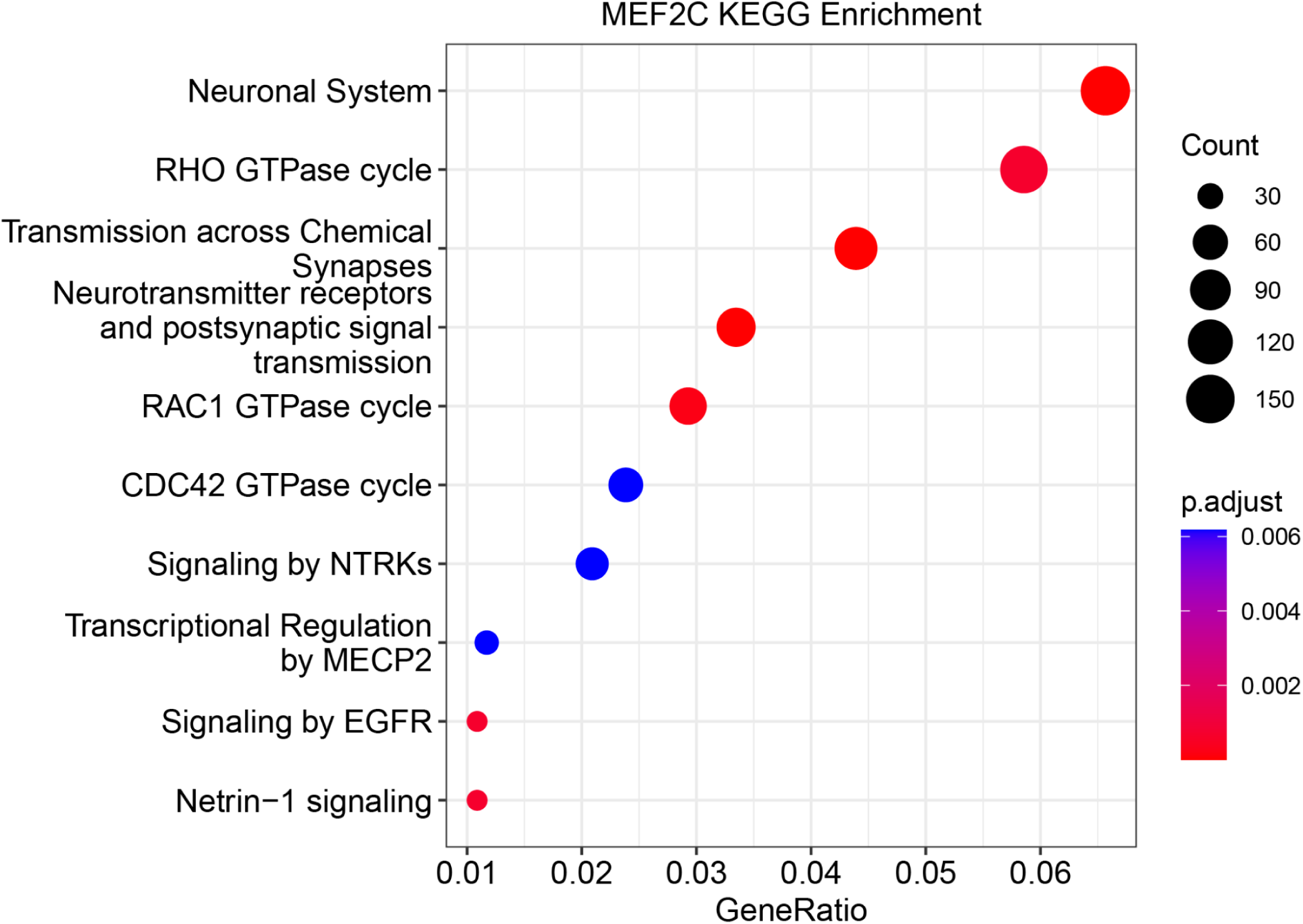
KEGG mapping of genes targeted by MEF2C in human prefrontal cortex. The set of genes bound by MEF2C as observed in CUT&Tag assays in neuronal nuclei sampled from postmortem human prefrontal cortex using fluorescence activated nuclear sorting maps to KEGG modules relevant to neuronal function, supporting the specificity of the experiment.

**Extended Data Fig. 6b.**
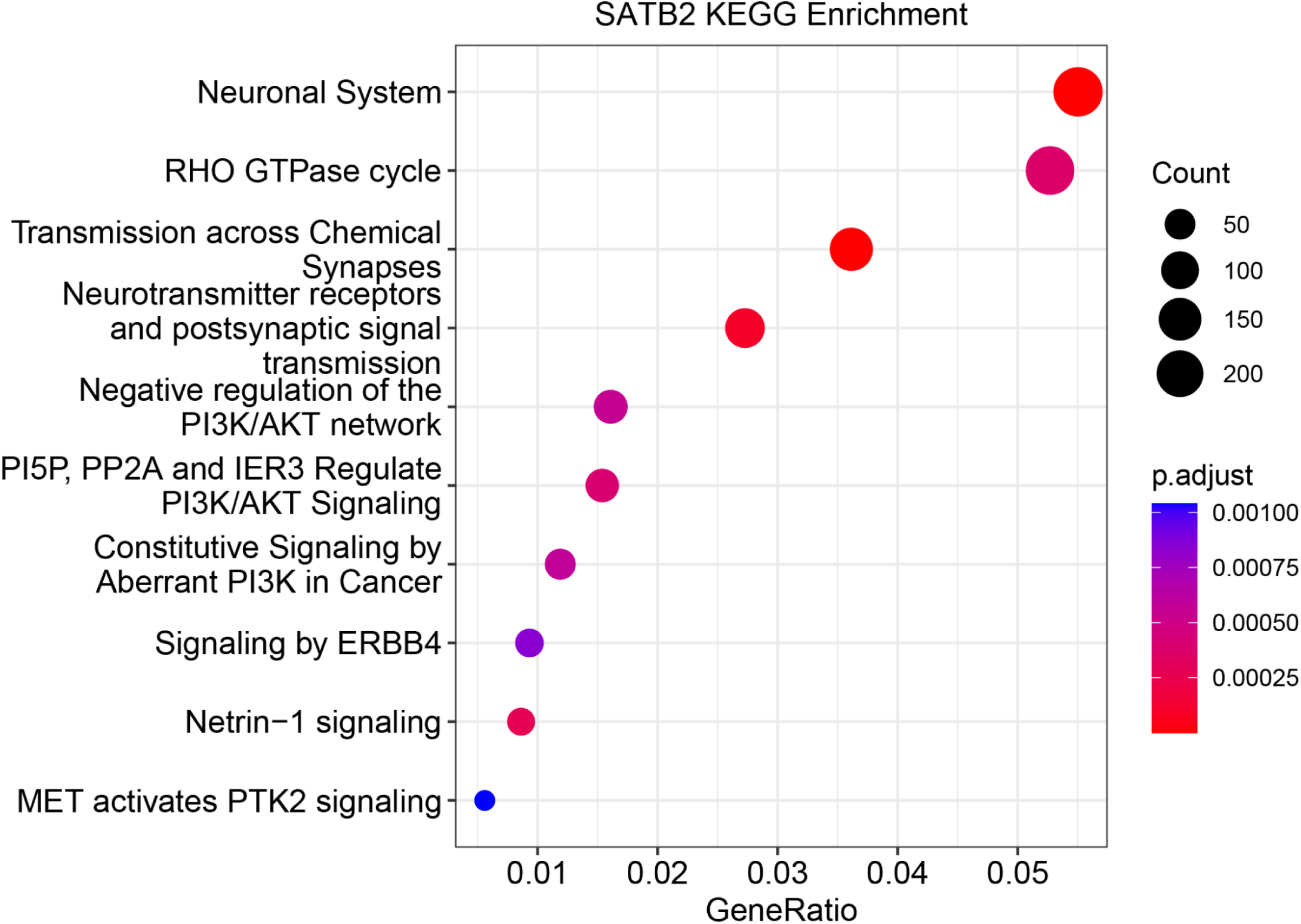
KEGG mapping of genes targeted by SATB2 in human prefrontal cortex. The set of genes bound by SATB2 as observed in CUT&Tag assays in neuronal nuclei sampled from postmortem human prefrontal cortex using fluorescence activated nuclear sorting maps to KEGG modules relevant to neuronal function, supporting the specificity of the experiment.

**Extended Data Fig. 6c.**
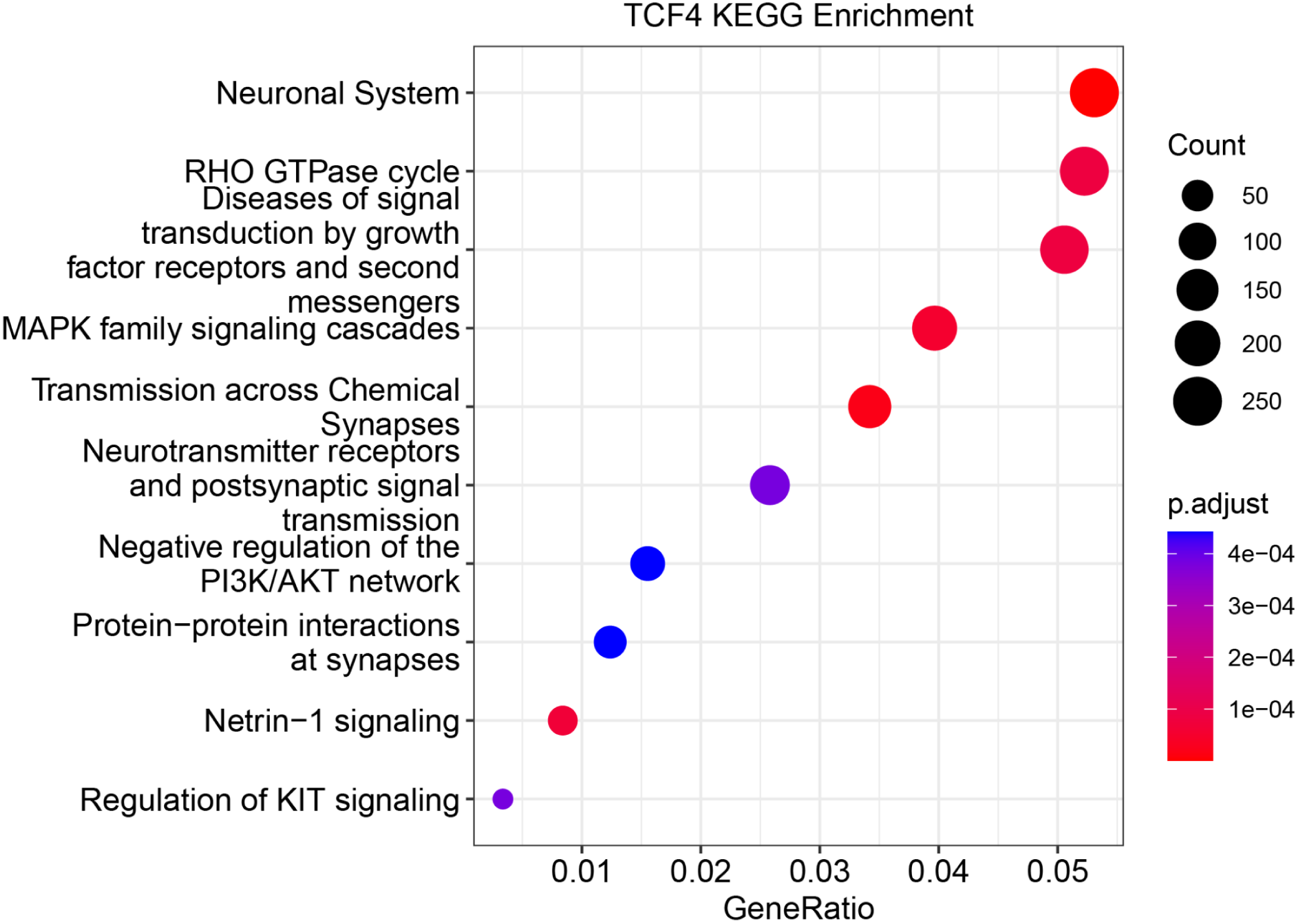
KEGG mapping of genes targeted by TCF4 in human prefrontal cortex. The set of genes bound by TCF4 as observed in CUT&Tag assays in neuronal nuclei sampled from postmortem human prefrontal cortex using fluorescence activated nuclear sorting maps to KEGG modules relevant to neuronal function, supporting the specificity of the experiment.

**Extended Data Fig. 7.**
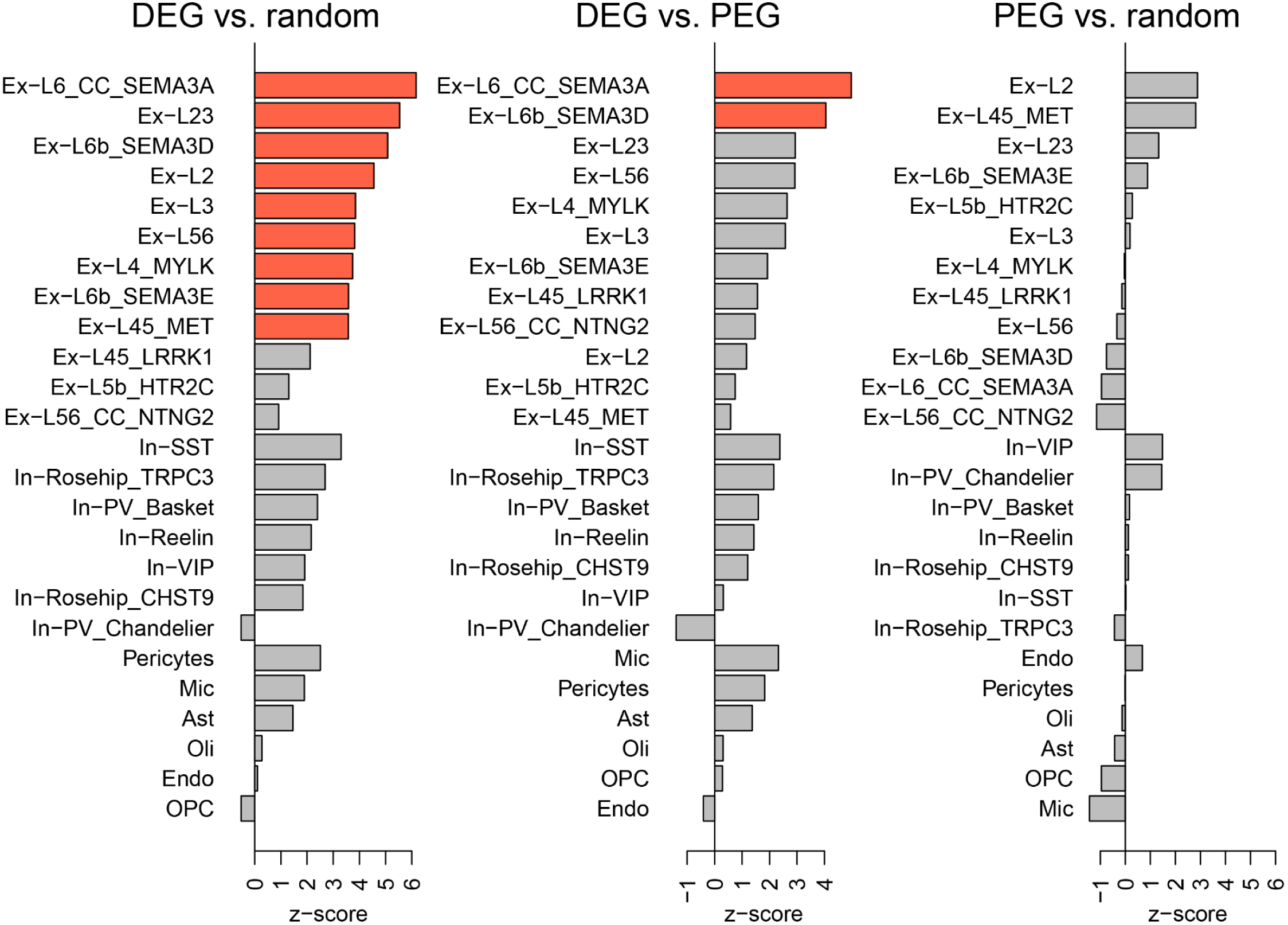
Schizophrenia cell-type specific DEG sets are enriched for GWAS association. Comparison of the level of enrichment of GWAS signal within each cell-type’s set of DEGs and an equal number of that cell type’s top preferentially expressed genes, as compared to an equal number of randomly selected genes computed by permutation test. Bars in red indicate that comparison’s Bonferroni corrected p>0.01.

**Extended Data Fig. 8.**
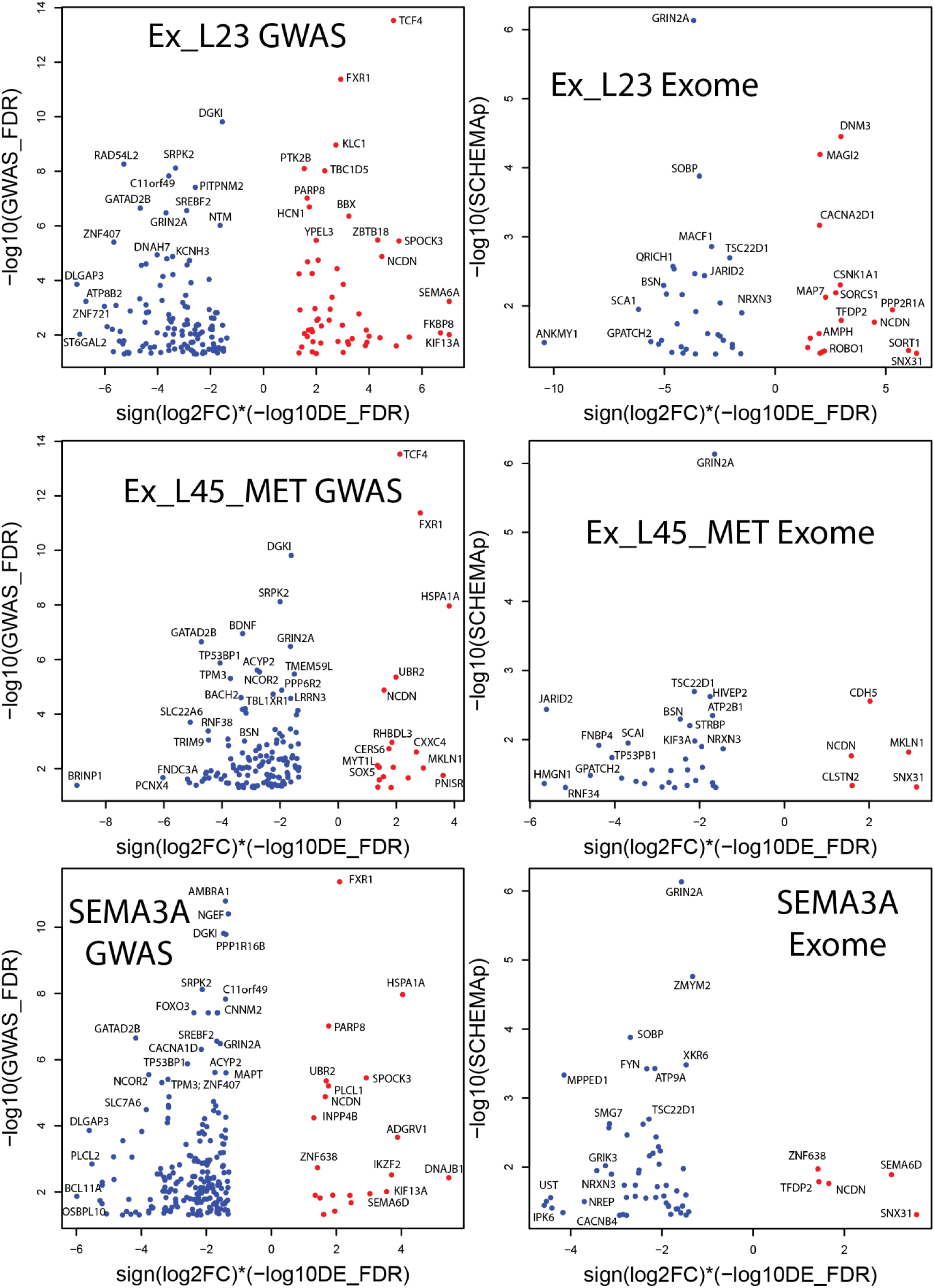
Gene level association between schizophrenia GWAS association and cell-type specific differential expression. Visualization of genes most strongly associated (FDR < 0.05) with common schizophrenia risk variants (GWAS) computed with H-MAGMA (y-axis, left) or rare schizophrenia risk variants (exome sequencing, y-axis, right) with significant expression changes in schizophrenia (x-axis) within additional populations of excitatory neurons.

**Extended Data Fig. 9.**
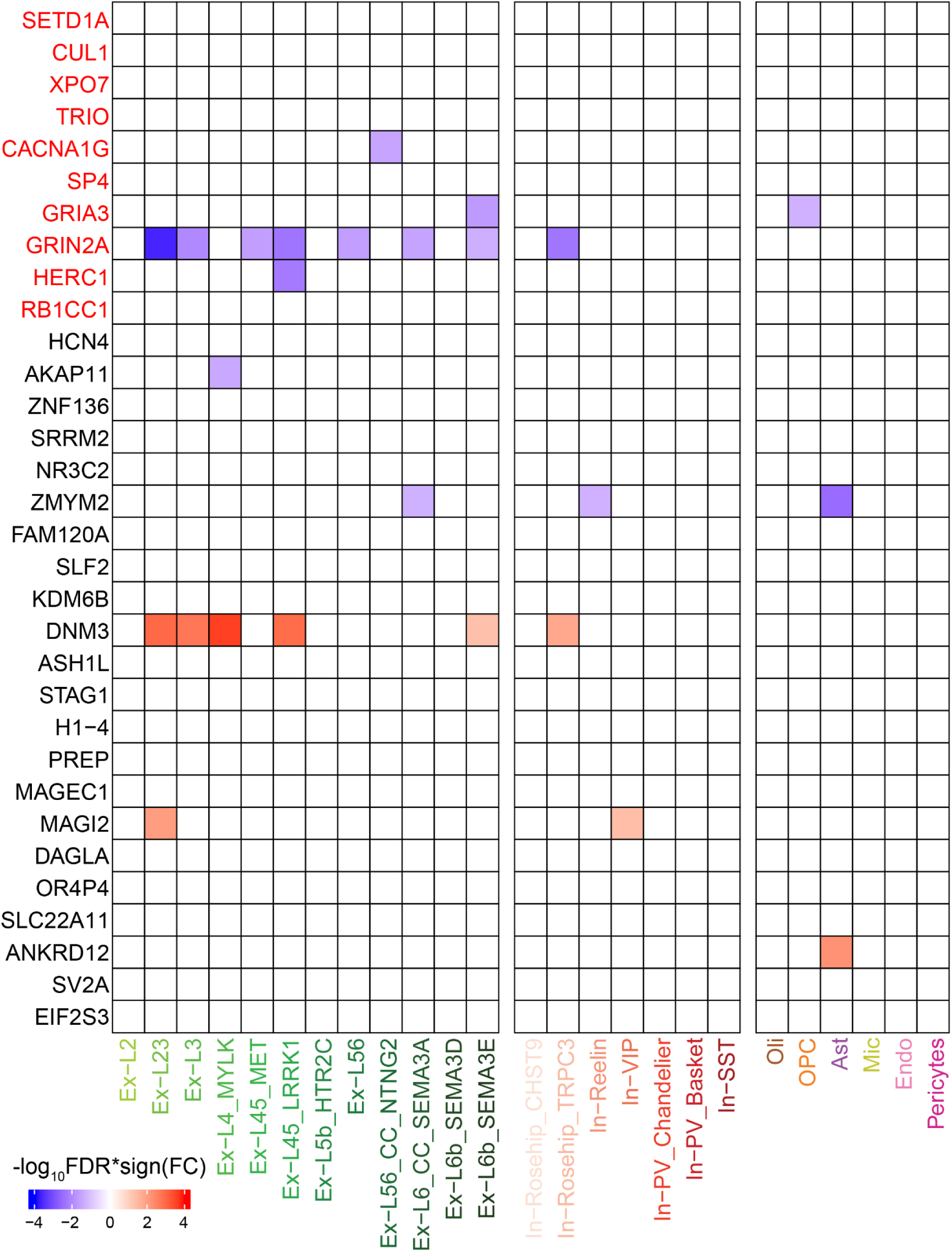
Cell-type-specific dysregulation of genes associated with rare protein coding variants in schizophrenia. Shown are the schizophrenia differential expression profiles for the 32 genes harboring rare protein coding variants associated with schizophrenia at FDR < 0.05. Schizophrenia association with genes named in red is exome-wide significant (71).

**Extended Data Fig. 10.**
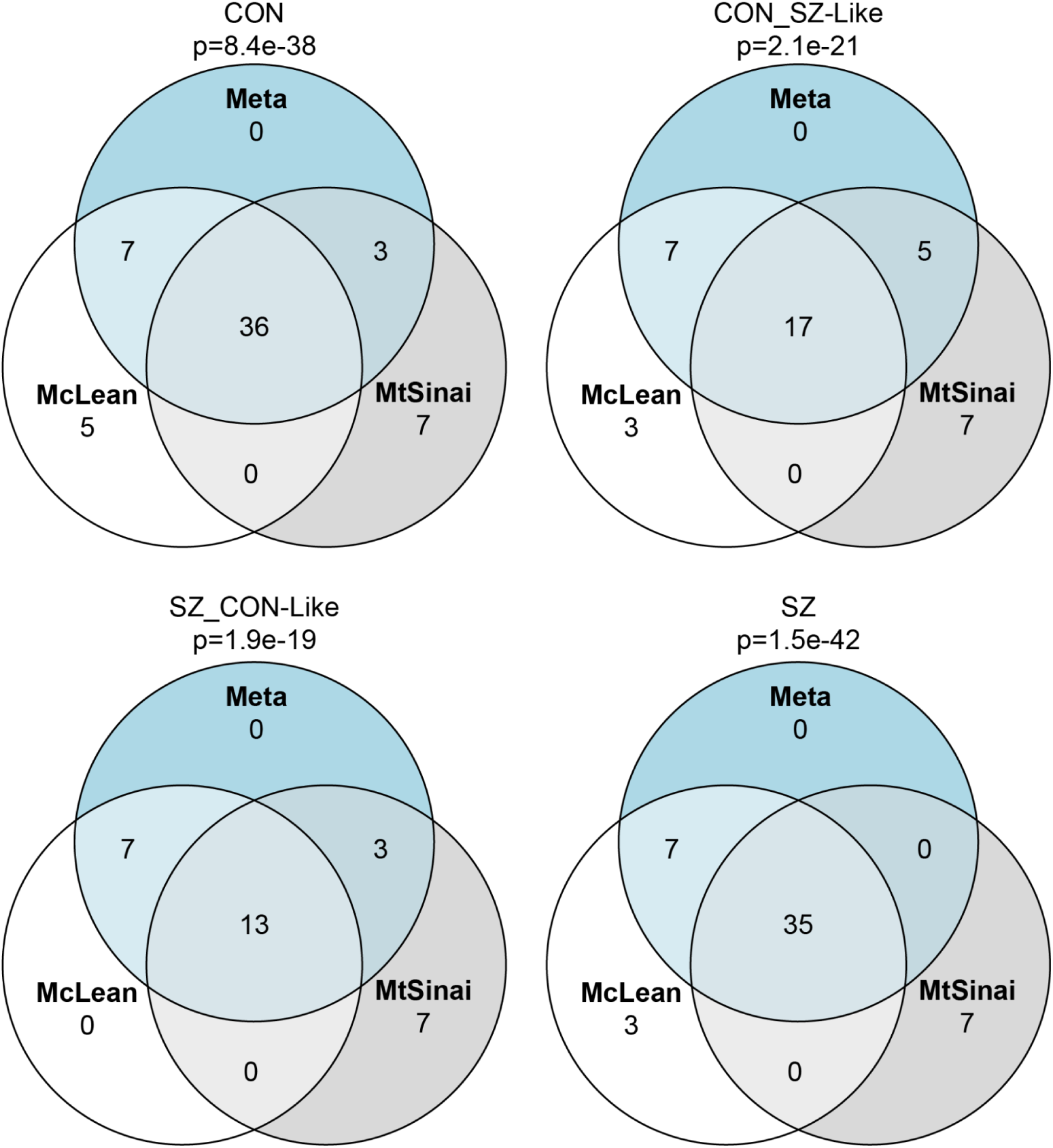
Consistency of diagnostic subgroup assignment across cohorts. Venn diagrams depict the overlap of subjects assigned to each diagnostic subgroup based on schizophrenia versus control gene expression changes observed within the McL cohort, the MSSM cohort, or the meta analysis results, demonstrating a high level of reproducibility across cohorts and analyses. Significance of multi-set intersections was calculated using the R software package SuperExactTest (132).

**Extended Data Fig. 11.**
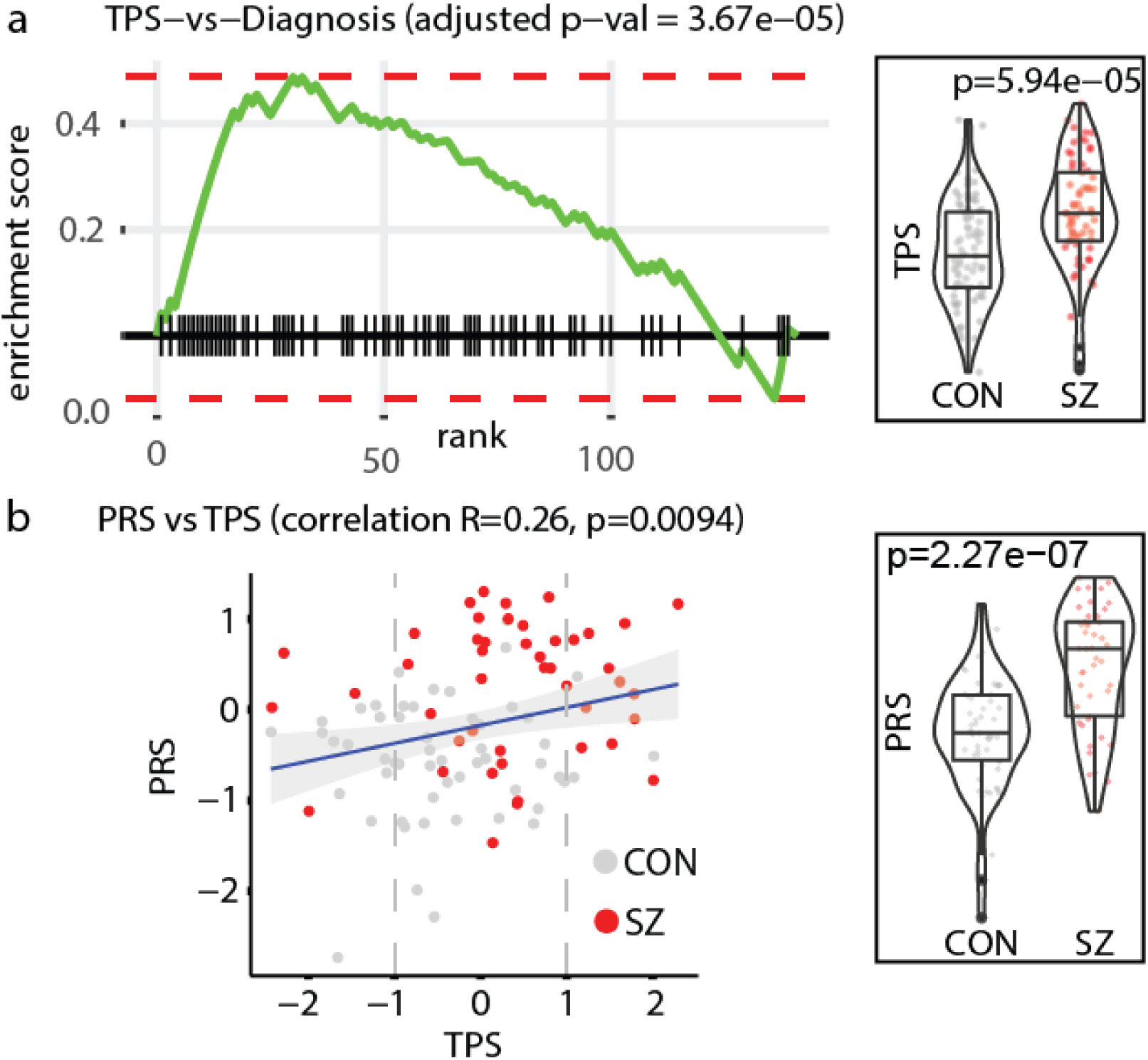
Transcriptional pathology score (TPS) is significantly associated with diagnosis (top) and with polygenic risk score (bottom), and both are significantly different between the control and schizophrenia groups.

**Extended Data Fig. 12.**
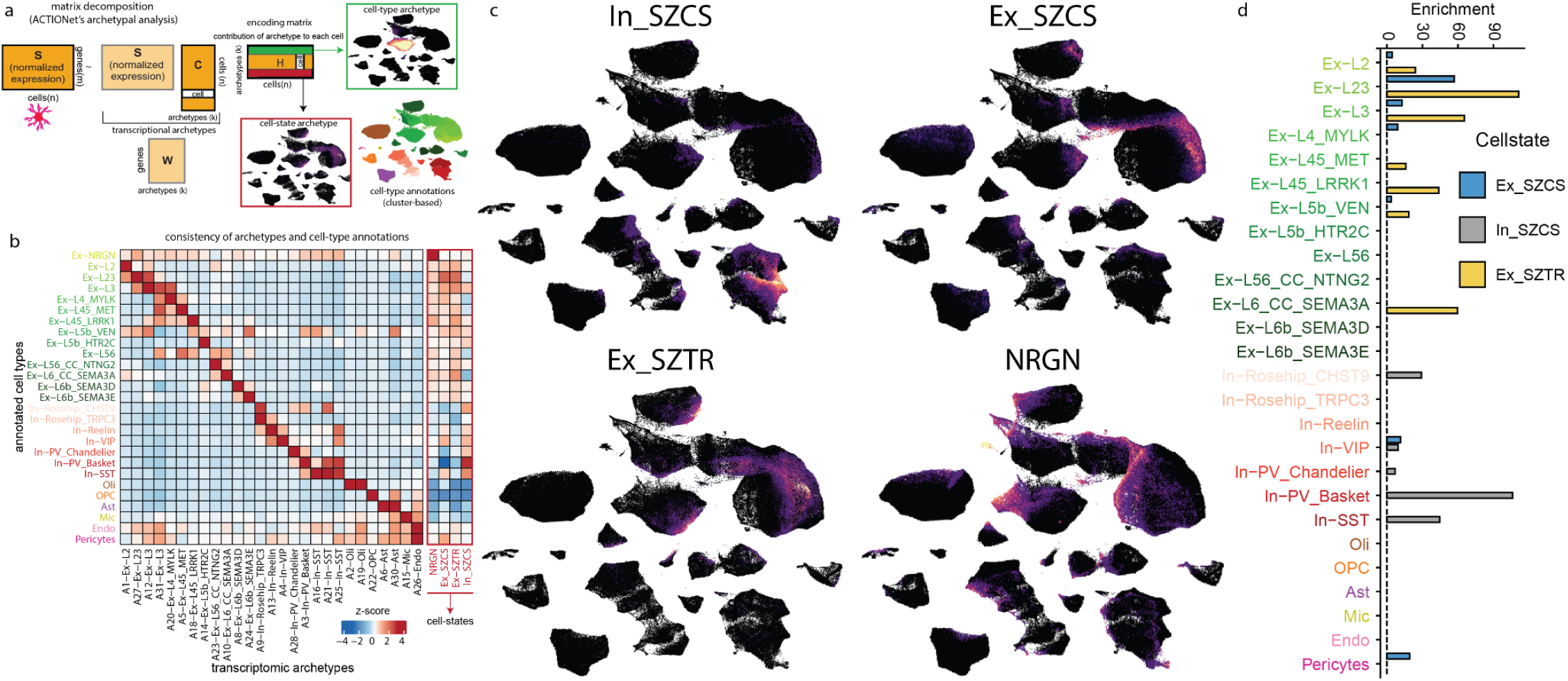
Cellular states identified by decomposition-based discovery of transcriptional signatures across all subjects and cells. a. Graphical representation of the matrix decomposition strategy used to identify transcriptional archetypes, most of which recapitulate cell-types, and a subset of which identify continuous cell-states spanning multiple cell-types. b. Association of all identified transcriptional archetypes with the 27 cell-types identified in our cluster based cell annotation, demonstrating that the majority correspond predominantly to a single cell-type, while four are associated with cells across multiple underlying cell-types. c. These four transcriptional signatures are expressed by cells across multiple neuronal populations, identifying transcriptionally defined cellular states. In_SZCS is most strongly expressed by inhibitory neurons, while the remaining three are most strongly expressed by excitatory neurons. The NRGN cellular state, marked by high expression of the *NRGN* gene, has been reported in prior studies of snRNA-seq in postmortem human brain(30). Cells with greater enrichment for the indicated cellular state’s transcriptional signature are more brightly colored. d. Enrichment of cell states within annotated cell types assessed by t-test and reported as t-statistics.

**Extended Data Fig. 13.**
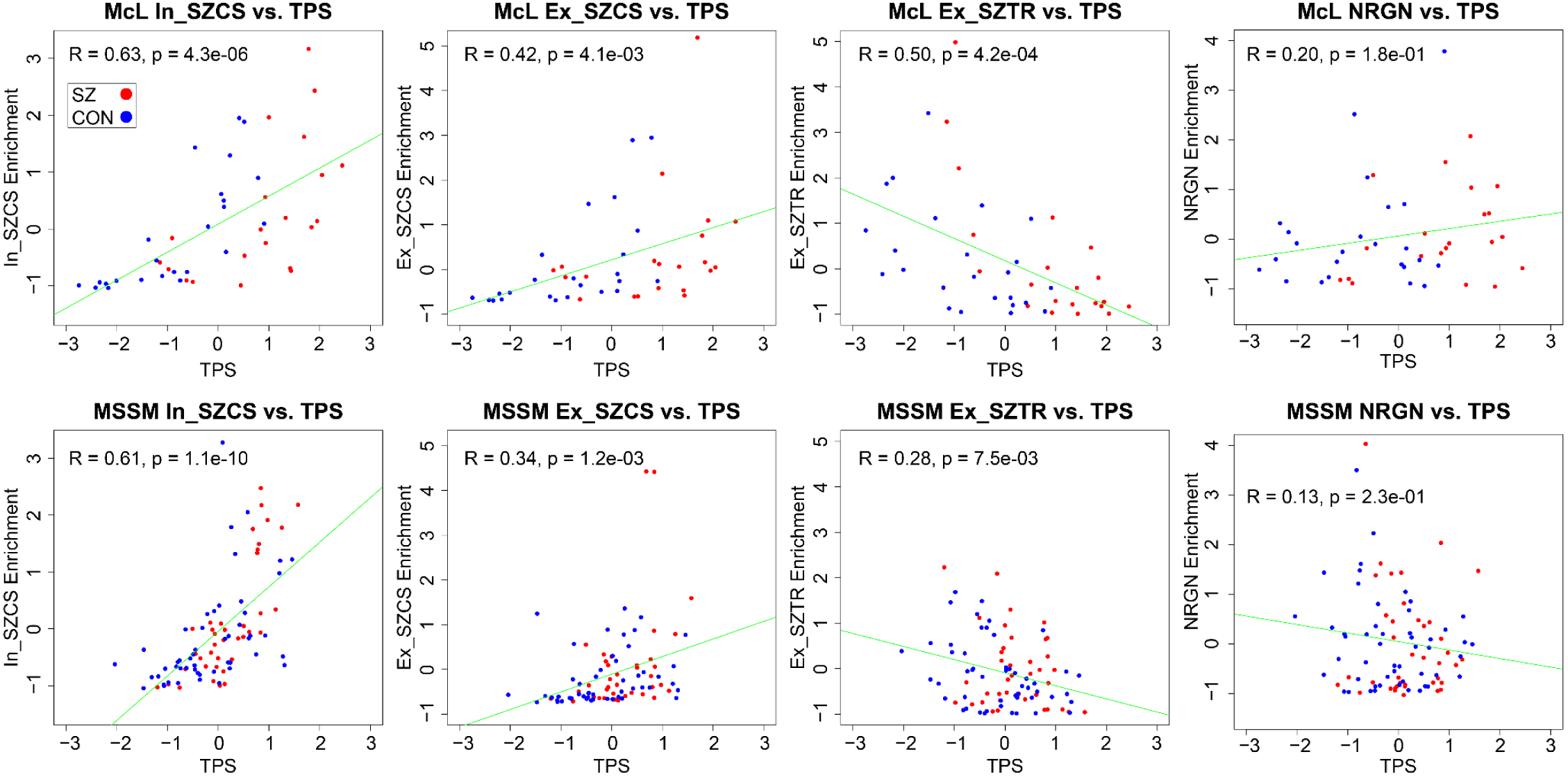
Consistency of cellular state enrichment with schizophrenia transcriptional pathology across cohorts. All three of the novel cellular states identified in this analysis show robust association with schizophrenia transcriptional pathology (as assessed across the complete cohort) within the assembled cohort (**Fig. 7d**) and each cohort independently. The previously described NRGN cellular state does not show this schizophrenia transcriptional pathology association.

**Extended Data Fig. 14.**
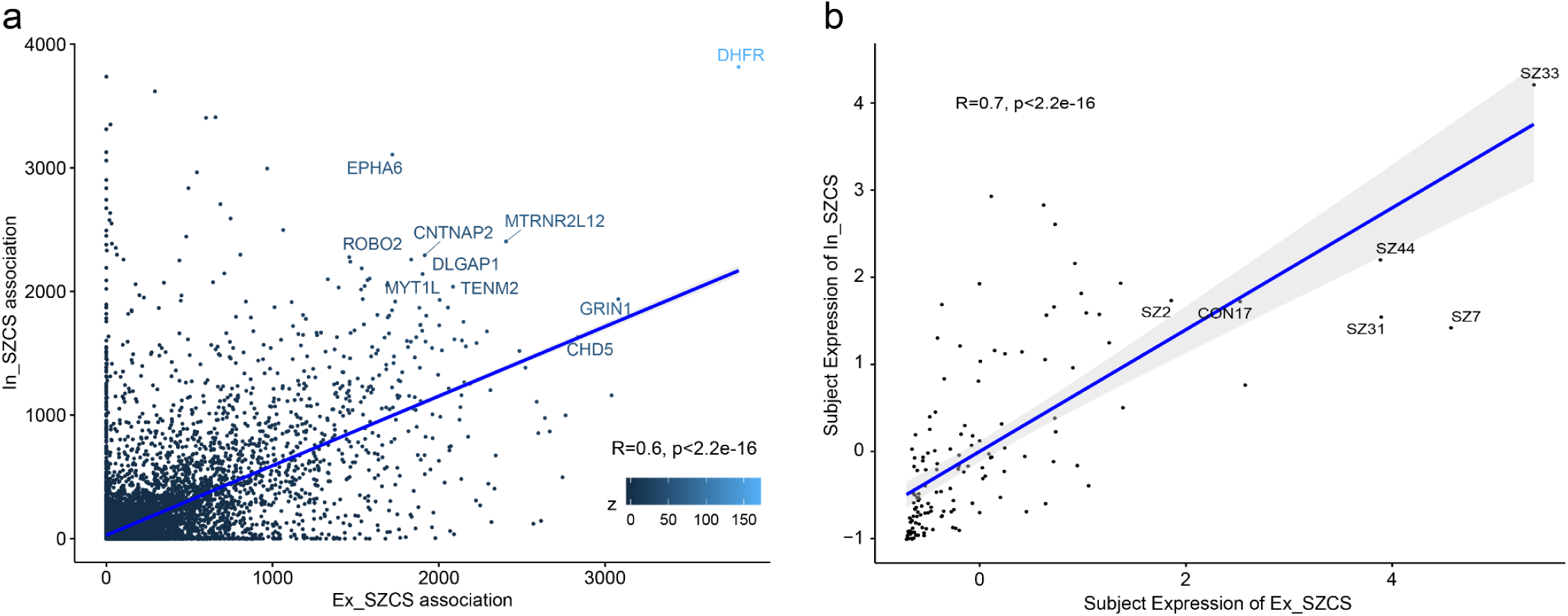
In_SZCS and Ex_SZCS are characterized by highly overlapping gene sets and are expressed by highly overlapping sets of subjects. a. Gene specificity scores for In_SZCS and Ex_SZCS are highly correlated, and identify top genes marking both transcriptional signatures. b. Expression of In_SZCS and Ex_SZCS within subjects is highly correlated.

**Extended Data Fig. 15.**
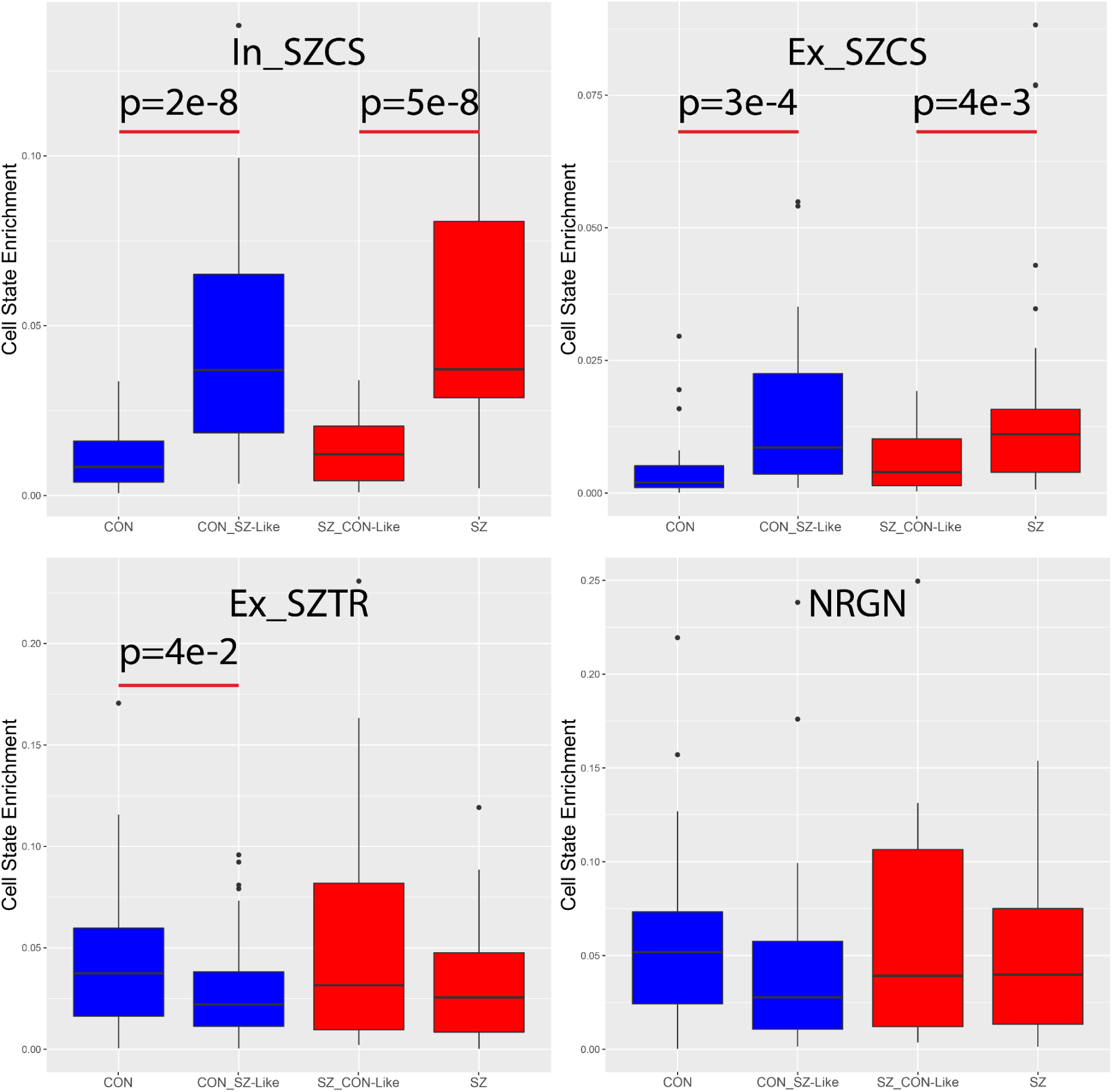
Enrichment of cellular states in subjects across diagnostic subgroups identified by subject transcriptional pathology score. Boxes represent the interquartile range with whiskers indicating the highest or lowest non-outlier values.

**Extended Data Fig. 16.**
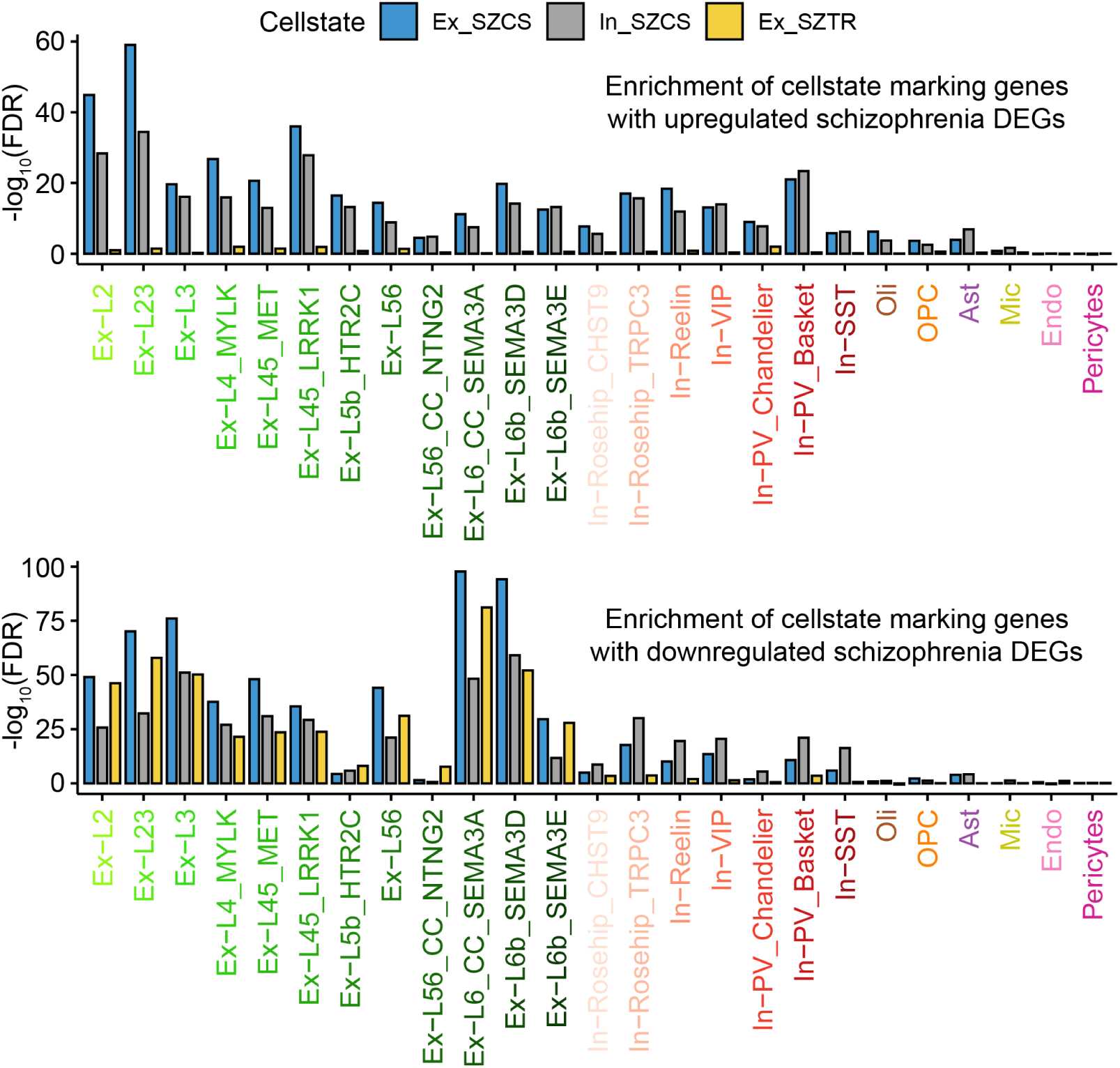
Gene Set Enrichment Analysis of each cell-type’s up and downregulated schizophrenia DEGs against all genes detected in the snRNA-seq dataset ranked by their association with the Ex_SZCS, In_SZCS, and EX_SZTR transcriptional signatures.

**Extended Data Fig. 17.**
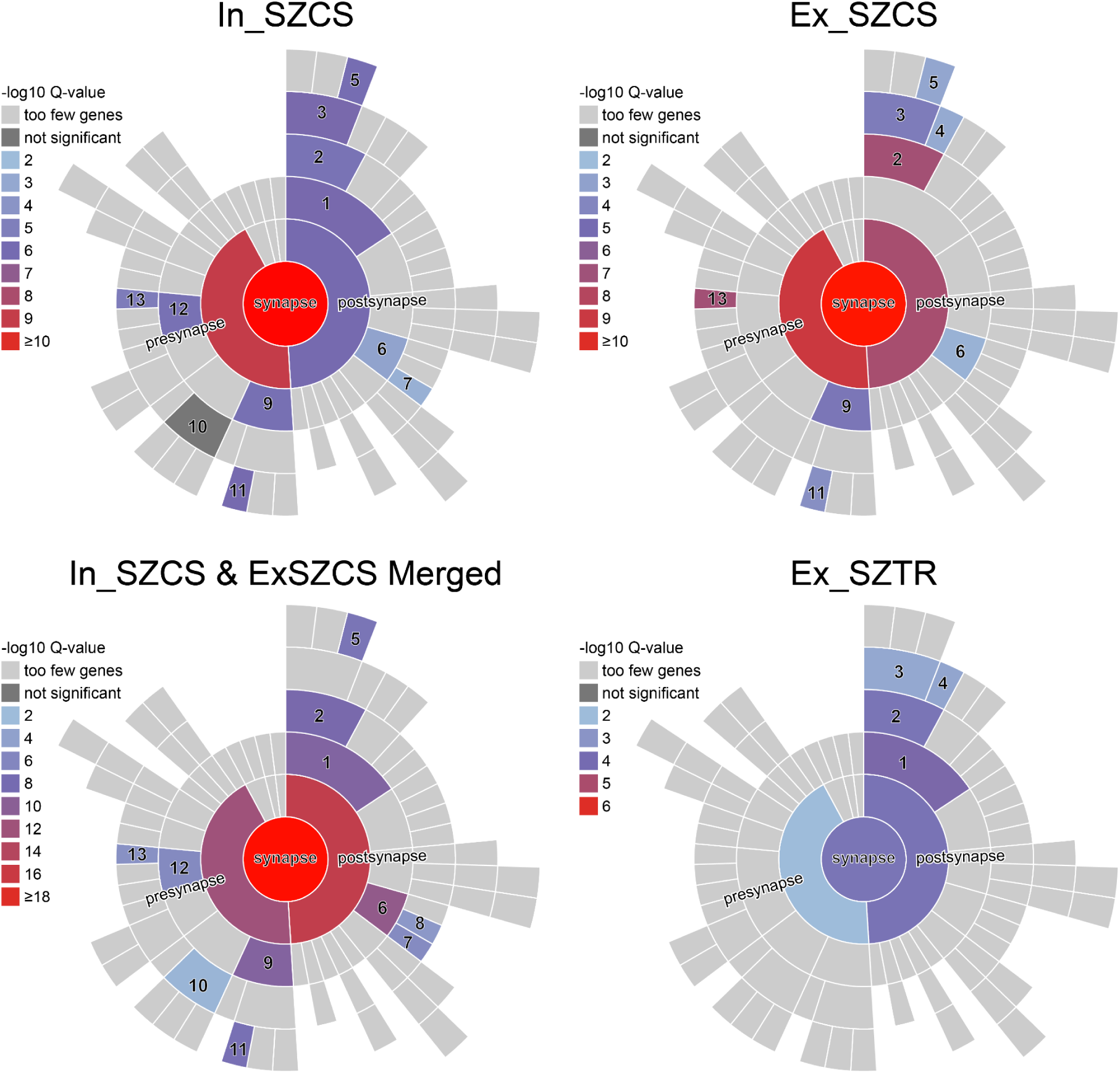
SynGO analysis of the 100 top genes marking each cellular state associated transcriptional signature. 1 - postsynaptic specialization; 2 - postsynaptic density; 3 - postsynaptic density membrane; 4 - postsynaptic density, intracellular component; 5 - integral component of postsynaptic density membrane; 6 - postsynaptic membrane; 7 - integral component of postsynaptic membrane; 8 - extrinsic component of postsynaptic membrane; 9 - presynaptic active zone; 10 - synaptic vesicle membrane; 11 - integral component of synaptic active zone membrane; 12 - presynaptic membrane; 13 - integral component of presynaptic membrane.

